# Ultra-efficient causal deep learning for Dynamic CSA-AKI Detection Using Minimal Variables

**DOI:** 10.1101/2023.12.04.23299332

**Authors:** Qin Zhong, Yuxiao Cheng, Zongren Li, Dongjin Wang, Chongyou Rao, Yi Jiang, Lianglong Li, Ziqian Wang, Pan Liu, Yawei Zhao, Pei Li, Jinli Suo, Qionghai Dai, Kunlun He

## Abstract

Cardiac surgery-associated Acute Kidney Injury (CSA-AKI) is a significant complication that often leads to increased morbidity and mortality. Effective CSA-AKI management relies on timely diagnosis and interventions. However, many cases of CSA-AKI are detected too late. Despite the efforts of novel biomarkers and data-driven predictive models, their limited discriminative and generalization capabilities along with stringent application requirements pose challenges for clinical use. Here we incorporate a causal deep learning approach that combines the universal approximation abilities of neural networks with causal discovery to develop REACT, a reliable and generalizable model to predict a patient’s risk of developing CSA-AKI within the next 48 hours. REACT was developed using 21.5 billion time-stamped medical records from two large hospitals covering 23,933 patients and validated in three independent centers covering 30,963 patients. By analyzing the causal relationships buried in the time dimensions, REACT distilled the complex temporal dynamics among variables into six minimal causal inputs and achieved an average AUROC of 0.93 (ranging from 0.89 to 0.96 among different CSA-AKI stages), surpassing state-of-the-art models that depend on more complex variables. This approach accurately predicted 97% of CSA-AKI events within 48 hours for all prediction windows, maintaining a ratio of 2 false alerts for every true alert, improving practical feasibility. Compared to guideline-recommended pathways, REACT detected CSA-AKI on average 16.35 hours earlier in external tests. In addition, we have established a publicly accessible website and performed prospective validation on 754 patients across two centers, achieving high accuracy. Our study holds substantial promise in enhancing early detection and preserving critical intervention windows for clinicians.

## Main

Annually, over two million patients globally undergo cardiac surgery to treat valvular heart disease or to correct congenital heart defects^1,2^.Despite advancements reducing procedure mortality, the invasive nature of these surgeries—marked by extensive incisions and complex, lengthy operations—often results in significant post-operative complications, with the highest incidence of cardiac surgery-associated acute kidney injury (CSA-AKI)^3^. Those with severe CSA-AKI have a mortality rate 3-8 times higher than others, and even with in-hospital renal recovery, CSA-AKI significantly independently associated their 10-year mortality risk^1^. However, the prognosis for CSA-AKI patients can be improved when timely interventions are applied, such as hemodynamic stabilization and volume optimization^4,5^. But there’s a catch: the intervention windows for effective intervention is limited, making early detection crucial.

In clinical practice, the diagnosis of acute kidney injury (AKI) often hinges on creatinine levels. However, kidney damage can manifest before a significant rise in creatinine^6^. Emerging biomarkers promise earlier detection, but their adoption is constrained by modest discrimination capabilities, the invasive and costly nature of the associated diagnostic procedures^7,8^. The Cleveland score^9^ and Mehta score^10^, based on traditional statistical methodologies, remain the primary prognostic instrument in contemporary clinical practice due to its inherent simplicity, methodological transparency, and ease of implementation have solidified its position. However, its reliance on a static analytical framework limits its predictive accuracy in the dynamically evolving clinical scenario. This deficiency is particularly evident in the cardiac surgical cohort, characterized by extended median hospitalizations—15 days on average—where patients’ clinical trajectories may undergo swift and unpredictable alterations^11^.The issue is exacerbated by the complexity of metabolism and individual variations in drug response, which obscure the identification of optimal intervention windows for CSA-AKI, often resulting in patients observed as at severe stages.

Neural networks, renowned for their proficiency in handling high-dimensional and time-series data, demonstrate potential in dynamically tracking patient conditions^12,13^. However, while adept at identifying intricate relationships, neural networks often conflate correlation with causation among variables, raising concerns about the reliability of their diagnostic outcomes and potentially resulting in suboptimal or dangerous decisions^14–16^. Furthermore, this inherent complexity, heightened by sensitivity to input data, impedes its broad applicability in clinical contexts. Additionally, artificial intelligence (AI)’s dependence on ultra-high-dimensional features during prediction muddies the path from data to actionable evidence. For instance, Tomasev et al.^17^ introduced a model utilizing 620,000 entries from hundreds of features for real-time AKI prediction. Despite its impressive performance, this approach necessitates collecting an extensive number of inputs in any prospective study to ensure accurate predictions. In practical clinical use, the absence of any single input could compromise or even incapacitate the predictive algorithm^18^. These highlight a clinical conundrum in neural networks application: the very above attributes that render AI invaluable for data analysis simultaneously make it vulnerable from a statistical viewpoint^14,19^.

Bridging the gap between traditional statistical inference and advanced predictive capabilities, causal machine learning emerges as a critical intersection of AI and statistics, marking a shift from mere prediction to a more profound understanding^20,21^. To facilitate dynamic prediction of CSA-AKI, we have developed a temporal causal deep learning architecture tailored for medical data, which is named REACT (Real-time Evaluation and Anticipation with Causal disTillation). As demonstrated in Figure 1, REACT combines the universal approximation abilities of neural networks with a causal discovery module to eliminate confounders or spurious variables^22–27^. This approach, simulating the process of Randomized Controlled Trials (RCT), sequentially conducts simulated interventions on all variables to assess their causal effects on the outcomes. It generates causal graphs and builds neural networks based on these graphs, identifying interactions and spatiotemporal dynamics buried in large, time-varying datasets, thereby reducing the likelihood of data-driven errors in AI models. In addition, our model significantly lowers the number of input variables required during application, shifting computational intensity and complex variable input to the training phase. This represents a principal advantage over existing algorithms that rely heavily on high-dimensional data inputs.

**Figure 1.**
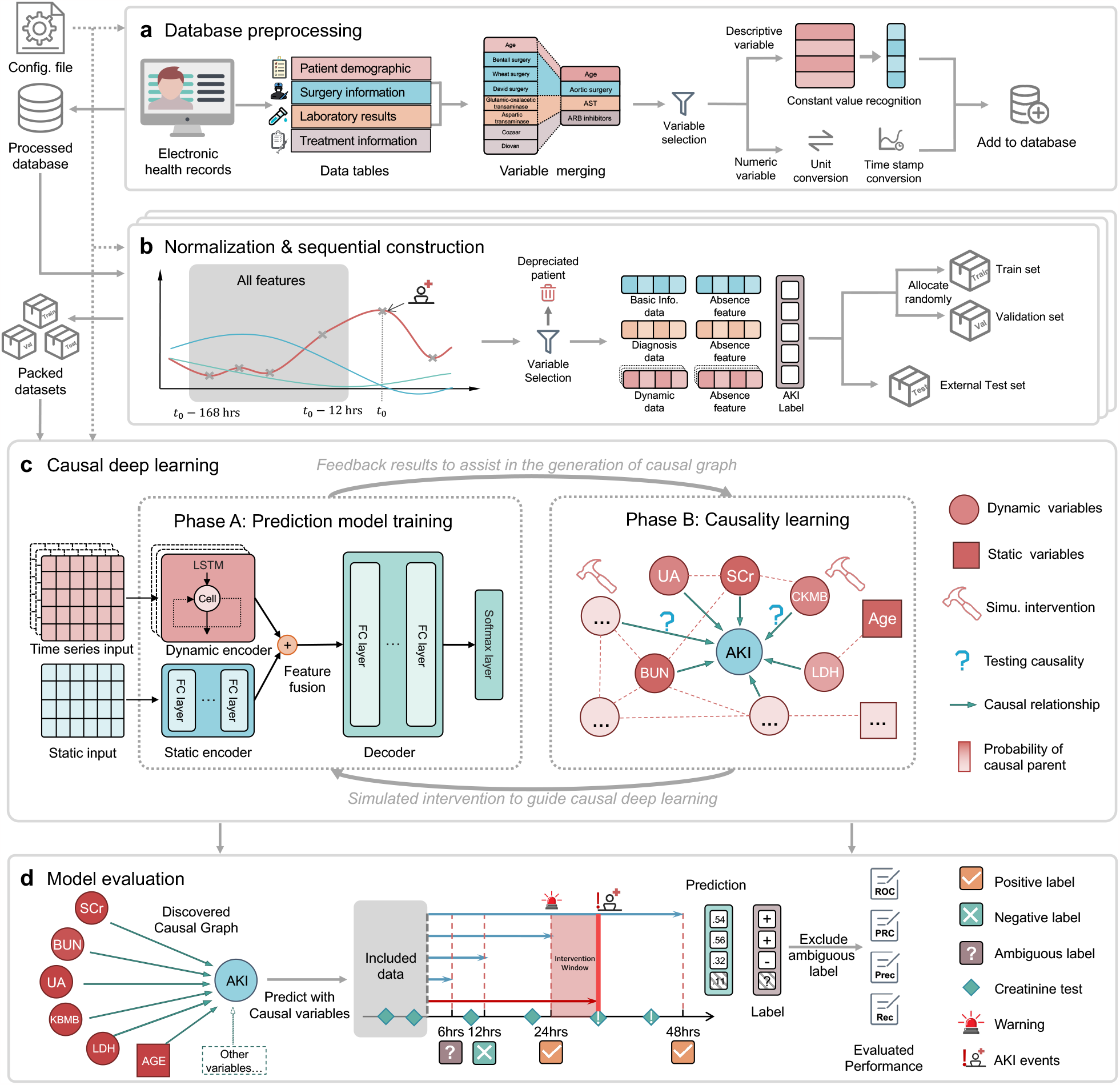
Overview of the model development. **a**, Database preprocessing. The raw database is collected from electronic health records of all patients with cardiac surgery and then processed by merging the same items with different names (such as laboratory tests, and vital signs) coding the same variable, excluding the rarely-present variables, converting units and time-stamps, excluding artifacts, and mapping textual test results to real-valued numbers. **b**, Normalization and sequential construction for each patient. The dynamic variables of each patient are extracted from the preprocessed database and then sampled to structured time-series with a two-hour time grid. The presence and absence of each variable are presented with a distinct feature. CSA-AKI labels are obtained at each time point of assessment with multiple prediction windows (i.e. CSA-AKI within 6, 12, 24, or 48 hours). All data and label pairs are divided into a training set, an internal (or in-distribution) validation set, and an external (or out-of-distribution) testing set. **c**, Causal deep learning. Our model consists of two iterative phases. The prediction model training phase predicts risks for CSA-AKI at each time-points. The Causality learning phase learns causal graphs with a fixed prediction model. Both phases are performed with the help of simulated intervention, which helps to build a causal prediction model that remains stable across environments. **d**, Model evaluation. Our method is evaluated with only causal features as input, (e.g., only six variables). AUROC, AUPRC along with other criteria are calculated by excluding ambiguous labels (that no serum creatinine tests are within the corresponding prediction window).

In our model, we used 21.5 billion timestamped medical records as initial inputs, drawn from two large hospitals, encompassing 23,933 cardiac surgery patients. Through causal deep learning, we distilled six dynamic variables (most common and cost-effective) for actual application input and validation on three independent hospitals (30,963 patients). As causal relationships remain stable across different environments, predicting CSA-AKI using only causal variables improves our model’s universal applicability. To advance this field, we have created a user-friendly publicly accessible website. Bridging the “last mile” from predictive modeling to application, we conducted prospective validation in 754 patients across two centers. The application of this technology in AKI is set to constitute a paradigm shift in nephrology telemetry and revolutionize the treatment of hospitalized patients, transitioning from reactive to proactive care.

## Results

### Comprehensive clinical dataset from five major hospitals

The study incorporated data from five large-scale general hospitals: The First and Third Medical Centers of the Chinese PLA General Hospital in Beijing, China, were used for the derivation of the predictive model and internal validation(from 2000 to 2021); For model external tests, we utilized the Sixth and Seventh Medical Centers of the Chinese PLA General Hospital, along with the Nanjing Drum Tower Hospital in Nanjing, China (from 2000 to 2023). Our cohort, comprising 54,896 patients, was selected from a pool of approximately 6.19 million individuals with 8.3 million visits treated at five major general hospitals. All five hospitals contributing to a collective database of over 21.5 billion time-stamped medical records. The PLA Hospital system serves as the key medical facility in northern China, while the Nanjing Drum Tower Hospital holds a comparable status in the south. Figure 2b illustrates the population density distribution across China, and Figure 2c represents the patient distribution within our study. As such, our research captures a substantial patient demographic from across China.

**Figure 2.**
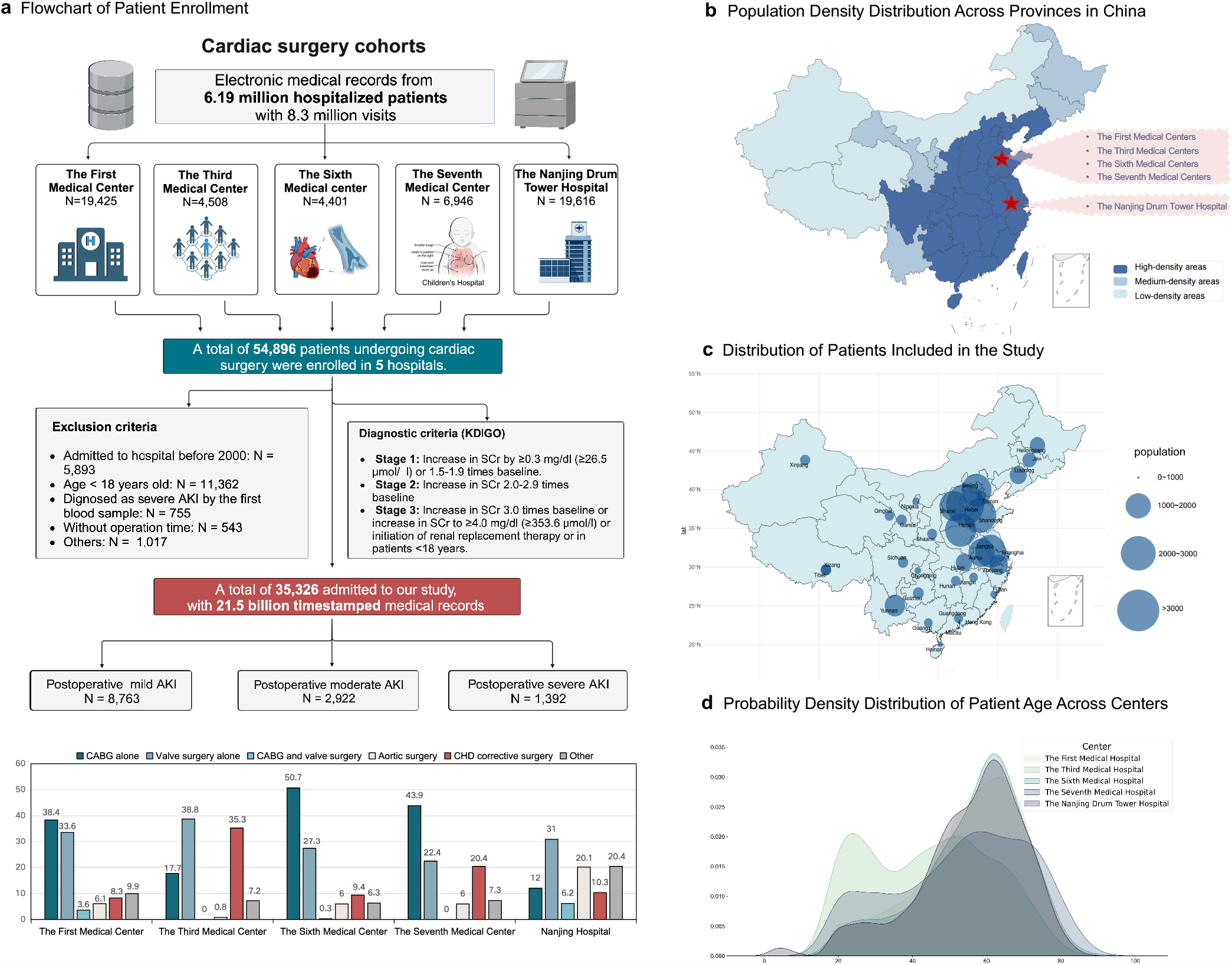
Comprehensive Visualization of Patient Demographics, Enrollment, and Treatment. **a**, Flowchart of Patient Enrollment: This diagram provides a detailed presentation of the criteria, number, and type of patients enrolled at each stage at the different medical centers, and also demonstrates the definitions of the different types of AKI. **b**, Population Density Distribution Across Provinces in China: This map shows the population density and geographical distribution of different medical centers, darker shades indicate higher density. **c**, Distribution of Patients Included in the Study: The size of the circle represents the number of patients enrolled in the study in their province. **d**, Probability Density Distribution of Patient Age Across Centers: The smooth lines represent the probability density function for each center’s patient age distribution.

After adhering to the exclusion criteria detailed in the Methods section, the derivation cohort included 14,513 patients (illustrated in Figure 2a), with a median age of 56.2 years and 28.1% being women. Baseline clinical characteristics and operative information for all participants are outlined in Figure 2a, 2d and *Supplements C*. In accordance with the modified KDIGO (Kidney Disease: Improving Global Outcomes) definition, the incidence of AKI was 20.1% (n = 2,913 patients). An interdisciplinary team harmonized synonyms and consolidated or substituted approximately 213 synonymous variables, employing medical guidelines and remote supervision through standardized entity libraries. A suite of techniques was employed for comprehensive data verification, including threshold checking, evaluation of linear relationships, and machine-learning-based outliers detection. Also, we did not include procedural factors that may be difficult to measure, such as aortic cross-clamp time. All these enhance the transferability and practicality of the model. After a manual approval process led by senior physicians, around 34,600 data entries were rectified.

To achieve dynamic predictions along the temporal dimension, the dataset was structured into a time-sliced format, segmented at 2-hour intervals. The outcome for each segment was labeled using the KDIGO criteria: segments were designated as “AKI Stage I” (Mild AKI) if they displayed an elevation in serum creatinine by ≥0.3 mg/dL over a span of 48 hours, as “AKI Stage II” (Moderate AKI) if serum creatinine doubled, and as “AKI Stage III” (Severe AKI) if serum creatinine tripled or reached ≥ 4.0 mg/dL with an acute increment of at least 0.5 mg/dL. However, a significant 29.3% of these time slices, due to the absence of adequate creatinine data, were classified as “Unknown”. Overall, we identified 380,563 slices with AKI events of 2,913 patients from 1,753,290 time-segments of 14,513 patients in the derivation cohort.

### Enhanced predictions through joint causal discovery and deep learning

It has been widely discussed in the literature that deep learning, when based on associations rather than causation, may lead to unstable predictions^16^ (see Method). To reveal stable underlying causal structures and achieve more precise and generalizable predictive outcomes, we propose a novel two-phase algorithmic structure. This framework harnesses the universal approximation capabilities of neural networks^28^ and incorporates a causal discovery method tailored for time-series data^23,25^. Figure 1c illustrates the “Prediction model training phase” and the “Causality learning phase” (with the detailed training process illustrated in Extended Figure 1 and detailed algorithm discussed in Methods), showing how the two phases were cohesively performed to complement and enhance each other’s performance. During the “Prediction model training phase”, our model forecasts the risk of future CSA-AKI at successive time points, and the “Causality learning phase” focuses on learning causal graphs with fixed weights for the neural network. These two phases synergistically evolve during the training process, the former leverages the network’s universal fitting capabilities to construct resilient and robust prediction models, while the latter particularly mimics Randomized Controlled Trials (RCTs), sequentially conducting simulated interventions on all variables, steering the prediction model to evaluate their impact on outcomes, and progressively distilling the causal variables and eliminating the spurious ones^25,29,30^. In Methods, we mathematically show how our approach recovers the actual causality. Through this collaborative integration, we developed the “REACT” model (Real-time Evaluation and Anticipation with Causal disTillation). REACT is engineered to dynamically generate the risk score of a patient developing AKI at any stage within the following 6, 12, 24, 48 hours and provide physicians with proper intervention opportunities.

### High performance in predicting CSA-AKI

As demonstrated in Figure 1, REACT can dynamically forecast the likelihood of patients progressing to different AKI stages within 48 hours following surgery. Specifically, the model’s predictive efficacy for severe AKI represented by the AU-ROC, varied from 0.949 to 0.972. The predictive performance heightened as the proximity to the event narrowed (6 hours: 0.972, 12 hours: 0.971, 24 hours: 0.969, 48 hours: 0.949). This temporal trend was consistently observed in predictions for AKI stages II and III, for detailed results, see Figure 3 and *Supplements F*. In scenarios featuring imbalanced datasets, the AUPRC provides a more nuanced assessment than the AUROC. For severe AKI predictions, our model achieved an AUPRC spanning 0.665 to 0.739 (as shown in *Supplements G*: 6 hours: 0.739, 12 hours: 0.737, 24 hours: 0.724, 48 hours: 0.665), Furthermore, REACT showed great accuracy for mild and moderate AKI (stages I and II) predictions within 48 hours, registering an AUROC between 0.93 and 0.95 and an AUPRC from 0.65 to 0.73. Overall, the model exhibited increased precision for more severe AKI predictions. Continuous scores adeptly approximate the time to event failure (as portrayed in Extended Figure 6). Following a post hoc recalibration using isotonic regression, we achieved near-perfect alignment between the model’s score and the observed risk.

**Figure 3.**
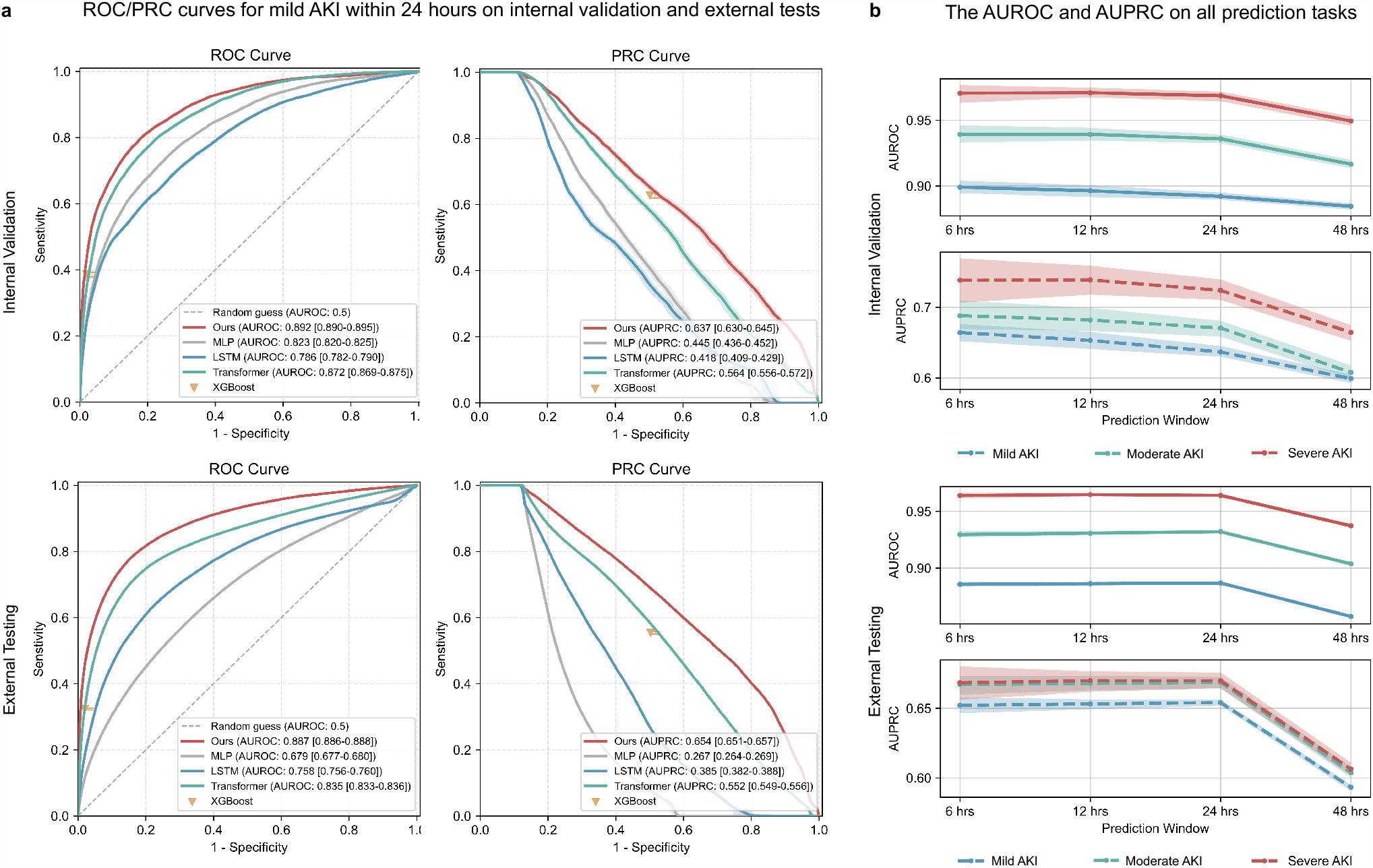
Model performance. **a**, ROC and PRC curves for mild AKI within 24 hours, on internal validation (randomly allocated) and external testing datasets (the sixth and the seventh medical center of Chinese PLA General Hospital, and Nanjing Drum Tower Hospital, which is not included in the training and validation datasets), comparing with baseline methods without casual deep learning (see methods), and the corresponding. **b**,The AUROC and AUPRC on other prediction tasks and windows. Our model supports multi-task learning, i.e., 4 kinds of prediction windows (6, 12, 24, 48 hours), each with 3 stages of AKI.

This was reflected with an overarching Brier score of 0.064. Calibration tests conducted across diverse patient subsets revealed that the model performs with consistent accuracy for most patient cohorts.

### Consistent efficacy across diverse patient cohorts

The performance of REACT was systematically evaluated across cohorts differentiated by age, gender, center, surgical type, year of admission and the mode of admission (details in Figure 4 and *Supplements H, I*). Overall, the model’s performance was positively correlated with the incidence rate of events and the completeness of data. The model showed exceptional efficacy in scenarios involving severe CSA-AKI, potentially due to more pronounced clinical indicators. More specifically, REACT consistently demonstrated better predictive performance for male patients compared to female patients across various tasks, with the AUPRC for males being approximately 1.16 to 1.34 times higher than that for females. This discrepancy might be related to the lower incidence rate of events in women. REACT’s predictive performance was relatively stable across different age groups, with the AUPRC generally exceeding 0.6. This stability was also observed across different types of surgeries; however, the model’s performance was less effective for multiple hybrid surgeries, underscoring the challenges these surgeries pose to predictions. The model’s performance was consistent across different admission years, but it was notably less effective for patients admitted after the year 2020. This difference may be attributable to the outbreak of the COVID-19 pandemic in 2019, which likely introduced unique medical dynamics affecting the model’s performance. Additionally, patients admitted through emergency departments had significantly higher event incidence rates and model performance compared to those with routine admissions, possibly due to more severe conditions and more frequent measurements of indicators.

**Figure 4.**
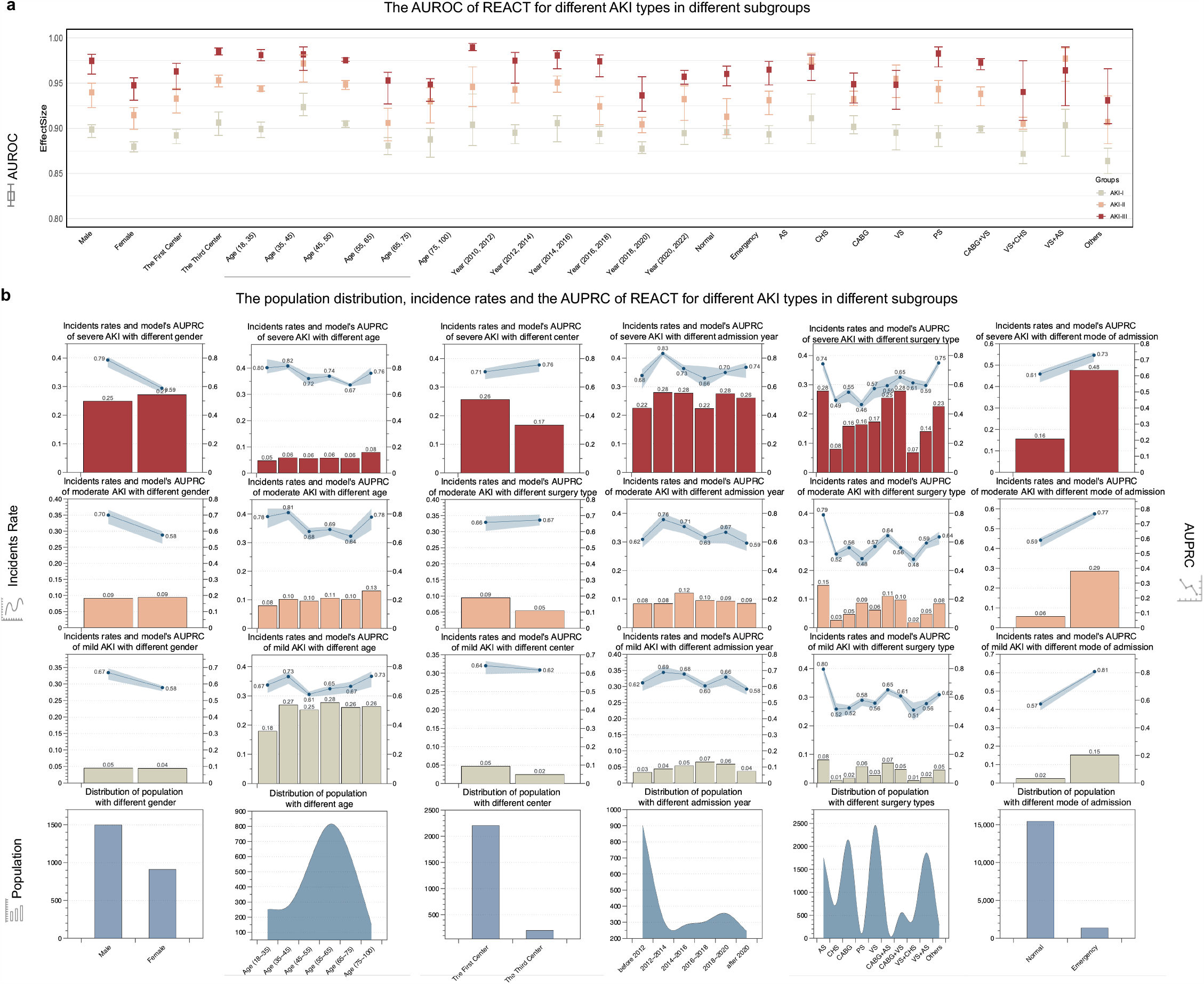
Model performance of different patient cohorts in internal validation datasets. **a**, Displays the AUROC predictions of REACT across all subgroups. Different colors signify the model’s performance at various CSA-AKI stages, with the vertical range indicating the model’s prediction performance variability within 48 hours of the event occurrence. The corresponding abbreviations in the operation type subgroup are as follows: AS, Aortic surgery; CHS, congenital heart disease corrective surgery; CABG, coronary artery bypass grafting; PC, pericardiectomy; VA, valve surgery. **b**, Illustrates the AUPRC predictions of REACT in different subgroups and provides detailed information about the population and incidence rate within each subgroup. Transverse subgraphs 1-5 specifically depict the model’s performance across varying surgical types, medical centers, age groups, admission years and genders. Longitudinal subgraphs 1-3 use bar graphs to show the incidence rate of CSA-AKI stage III-I among patients of different subgroups (as denoted on the left y-axis) and use plots to show the corresponding CSA-AKI prediction AUPRC with the shadow indicating performance variability within 48 hours of the event(as denoted on the right y-axis); In Longitudinal subgraph 4, the total number of patients in each subgroup is represented.

### The clinical approach of the REACT’s utilization

As a demonstration of how a hospital might employ REACT, we set a threshold to anticipate the event’s occurrence and recommend appropriate interventions with two false predictions for every true positive (details in *Supplements P*). This approach accurately predicts 97% of AKI events, on average 14.6 hours before any CSA-AKI event(internal validation). As the event approaches, the model’s predictive accuracy incrementally improves: 75.8% of cases are predicted successfully 48 hours in advance, and 85% are predicted 24 hours ahead. In scenarios involving patients with severe afflictions or those undergoing renal replacement, the model’s performance is notably enhanced, demonstrating a recall rate of 89% and a precision of 96%, as illustrated in Figure 3. We also provide a comprehensive report on the model’s recall rate, accuracy, precision, and other predictive metrics at different thresholds and time points in *Supplements M-O*. For the total inaccurate predictions(false-positives, detailed in Extended Figure 2). A deeper analysis revealed that 34.4% of these inaccuracies were due to delayed AKI onset within 24 hours, 8.2% are attributed to 24-48 hours after time of assessment. Only about half (50.7%) are actual false positives that didn’t reach the respective thresholds—a trade-off made to avoid alarm fatigue.

In our dataset, we observed that 40% of patients were already at AKI stages II-III when detected. As illustrated in *Supplements Q*, these patients had a relative risk (RR) of 2.38 (p *<*0.001) for mortality compared to those detected at stage I and experienced an average postoperative hospital stay extension of 1.14 day. This could be attributed to some patients missing the optimal intervention window, thus, early prediction for these patients becomes crucial. Our model achieved an 83% predictive accuracy within 24 hours for patients initially observed at stage I, extending the intervention window by 16.29 hours, which could significantly improve patient outcomes.

### Robust performance in external tests

The external testing set comprised 16,987 patients from three major medical institutions: the Sixth Medical Center, the Seventh Medical Center, and Nanjing Drum Tower Hospital (median age 60 years, 35% women). As illustrated in *Supplements C*, these three hospitals have notably diverse patient characteristics. Despite these variations, our model consistently outperformed other methods when applied to these datasets, achieving an average AUROC of 0.92. Similarly, calibration was good with Brier scores of 0.062(depicted in Figure 3, Extended Figure 6 and *Supplements J, K*). When applying the threshold determined from the training set, our model demonstrated average specificity rates of 0.81, 0.83, and 0.85 for mild, moderate, and severe CSA-AKI respectively, coupled with corresponding average recall rates of 0.78, 0.89, and 0.95. Notably, the model could predict 95% of events in advance, with an average lead time of 16.35 hours. An analysis of different subgroups within the external testing set yielded conclusions largely consistent with those from the training set(shown in Extended Figure 4). In addition, the model’s performance exhibited a more pronounced negative correlation with age and a more significant decline in performance post-2020. This decline could potentially be attributed to the unique medical dynamics introduced during the COVID-19 pandemic, as mentioned previously.

This aspect of the model’s performance, indicating a change in efficacy with patient age and external factors like the pandemic, underscores the need for ongoing model evaluation and adaptation in response to evolving clinical landscapes.

### Superior performance to traditional models

In this study, we conducted a comprehensive comparison of our model’s performance against widely recognized predictive algorithms in forecasting predefined outcomes across various timeframes. The comparative algorithms we selected include XGBoost^31^, a machine learning algorithm widely used in clinical settings due to its robustness and efficiency, the classic Multilayer perceptron (MLP)^28^, Long Short-Term Memory (LSTM)^32^, and Transformer models^33^, which demonstrate notable advantages in sequential data prediction for their revolutionary attention mechanism and have set new benchmarks in handling sequential data, particularly in complex tasks involving large datasets. Figure 3 and *Supplements F* illustrate the AU-ROC and AUPRC of each model over various time intervals, Our model outperformed the others at all time points. Taking the prediction of severe AKI within 24 hours as an example, REACT achieved AUROC scores of 0.969 and 0.964 in the internal validation set and external test set, respectively. These scores were significantly higher than those of XGBoost (0.65), MLP (0.913 and 0.739), LSTM (0.880 and 0.850), and Transformer (0.926 and 0.943). Notably, despite being based on entirely new data, REACT exhibited minimal performance fluctuation in the external test set, maintaining a median AU-ROC loss of just 0.008 (interquartile range 0.005–0.012). This performance markedly exceeded that of the comparator models (MLP: 0.151 0.144,0.163, LSTM: 0.027 0.020–0.030, and Transformer: 0.029 0.023,0.040). These findings unequivocally demonstrate the superior precision and stability of our React model in the dynamic prediction of CSA-AKI.

### Reliable predictions from minimal variables

Benefiting from the intrinsic relationships uncovered during the causal discovery phase, our finalized model requires only six indicators as input to outperform mainstream algorithms that rely on all variable inputs. It’s imperative to clarify that REACT’s predictive process, though based on only six causally significant indicators, does not imply that the model’s information is derived solely from these variables, i.e., our approach is not trivially performing feature selection before deep learning. The model comprehensively learns and captures the dynamic interplay among all variables within the temporal sequence and generates a causal graph to reveal the underlying causal structures, which serve as explanation results of the model. This enhances the model’s efficiency, generalizability, and applicability in real-world clinical settings. In the experiments shown in Extended Figure 5 we show that training a regular neural network (e.g., MLP, LSTM, Transformer) with only these six selected variables, i.e., performing feature selection before deep learning instead of our causal deep learning strategy, the AUROC/AUPRC scores are lower. Taking the prediction of severe CSA-AKI within 24 hours as an example, REACT achieved an AUPRC of 0.724 in the internal validation set, which is 1.18 to 3.20 times higher than algorithms trained using the selected causal variables. This superiority is even more pronounced in the external test set, where the improvement ranges from 1.10 to 7.44 times. These findings demonstrate that our causal deep learning approach not only increases generalizability by pinpointing causal variables but also prevents overfitting throughout the training process, thanks to a dropout-like^34^ sampling strategy (see Methods).

**Figure 5.**
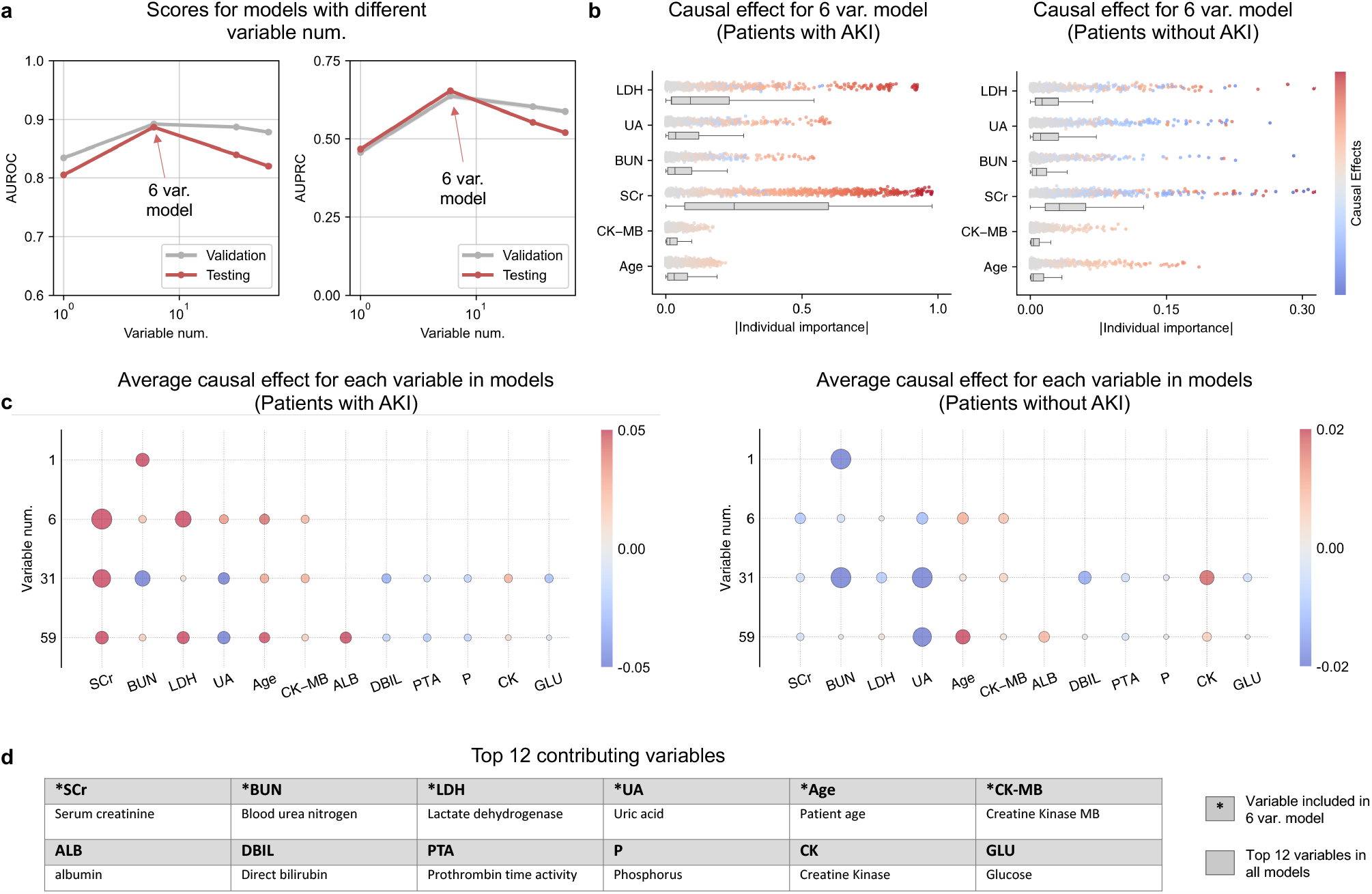
Importance analysis of predictive variables. **a**, The relationships between the number of causal variables, internal validation performance, and external testing performance. Here the baseline is Transformer. **b**, Local feature importance with the 6 variables setting calculated by analyzing individual direct causal effects. **c**, Global feature importance for various models with different causal variables calculated by analyzing direct causal effects. (show only the top 12 variables for simplicity). Y axis in (b) and x axis in (c) show absolute values of the importance, while the color demonstrates the signs. **d**, Full names of top 12 contributing variables.

**Figure 6.**
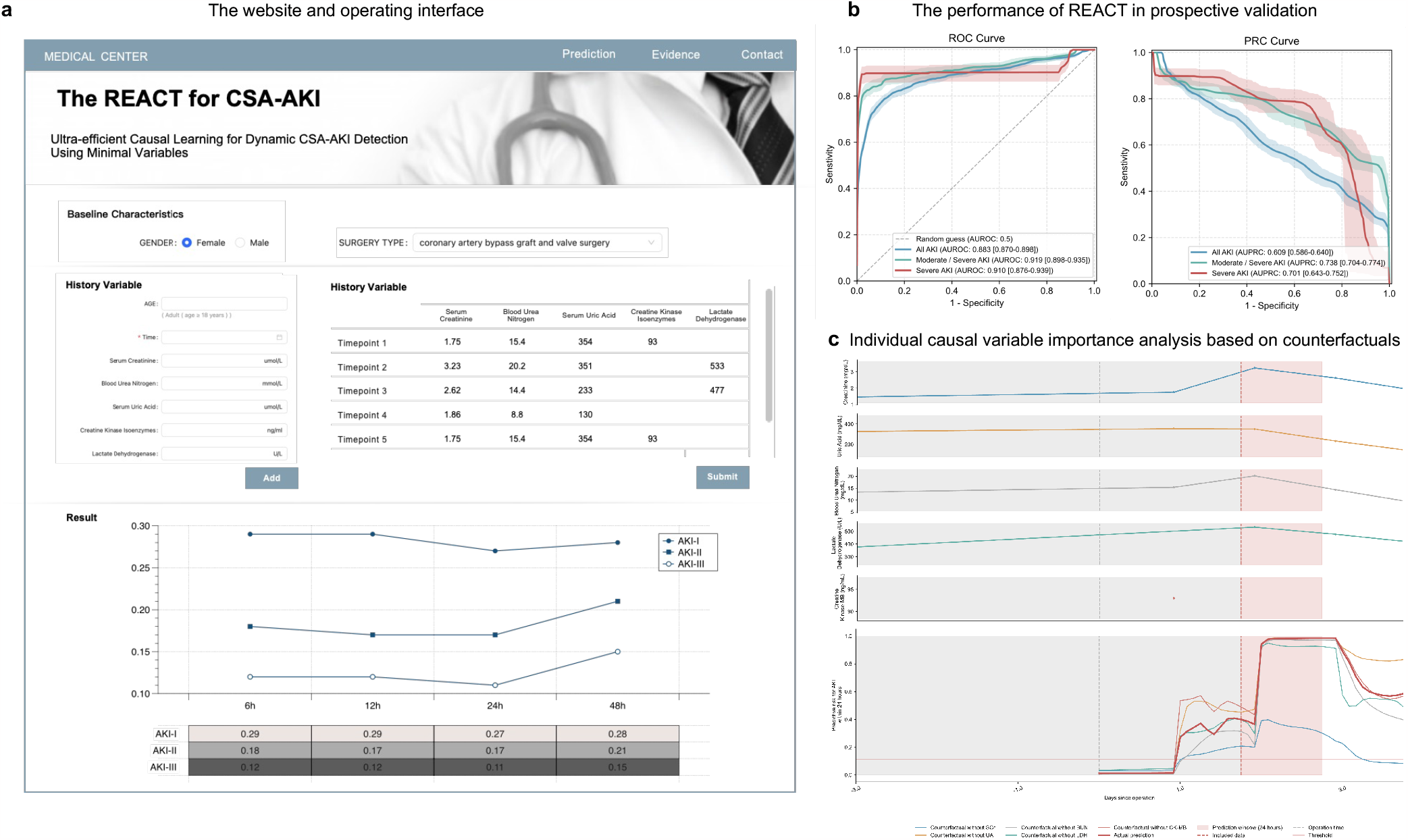
The Online platform and prospective validation. **a**, The website and operating interface. The platform allows for the input of baseline characteristics, including age, gender, and type of surgery. Users can also enter predictor variables such as time, and serum creatinine. The system is designed to accommodate multiple measurement data entries. Model output showcasing the risk score of AKI at different stages within the subsequent 48 hours. Detailed model output indicating moments when interventions are recommended. **b**, The performance of REACT in prospective validation. This subgraph presents ROC and PRC curves for all AKI stages within 24 hours. **c**, Individual causal variable importance analysis based on counterfactuals. A female patient in their 50s with hypertension, diabetes mellitus, and chronic heart failure was admitted to the hospital for coronary (aortic) coronary artery bypass grafting, mitral-valve replacement, and tricuspid valvuloplasty. The length of stay of the patients was 37 days. Creatinine, uric acid, and urea nitrogen indices were stable before surgery, with a slight increase in creatine kinase enzymes. Finally, the patient developed CSA-AKI.The individual importance by deleting one of the six variables (i.e., counterfactual questions “what if SCr / UA / … is another value?”, see Methods for details).

### Importance analysis of predictive variables based on causal inference

With the trained network and discovered causal graph, we can ascertain the contribution of each variable. This is achieved by comparing the predicted risk with its counterfactual version when absent—essentially gauging the shift in predicted probability upon altering a specific variable (details in Methods). A critical balance between model capacity and transferability is necessary when predicting CSA-AKI using causal variables. As shown in Figure 5c and 5d, the causal variables ultimately included in REACT are blood urea nitrogen, uric acid, Lactate Dehydrogenase, Creatine Kinase Isoenzyme, and age. Figure 5b illustrates their individual and collective importance. It is noteworthy that the model can still make predictions with a slight sacrifice in performance when a few variables are missing. We also provide detailed experimental data on the model’s performance with different numbers of causal variables in the *Supplements L* and considering the variability of available variables in actual clinical scenarios due to hospital differences, we also offer four versions of REACT with varying degrees of variable richness on our code repository (https://github.com/jarrycyx/UNN/tree/main/REACT).

### The website application and a prospective validation

To increase the accessibility for users and streamline the testing process of our model, we created a web-based platform (http://www.causal-cardiac.com) tailored for dynamic early alerts of CSA-AKI. As showcased in Figure 6a, the user interface provides the functionality to input baseline demographics and causal predictor variables with the capability for multiple data entries. Upon entering the required data, the underlying model, powered by a pre-trained neural network with integrated causal discovery, generates the likelihood of CSA-AKI episodes at multiple intervals over the next 48 hours. Importantly, the model indicates specific instances recommending either the initiation or escalation of medical interventions, thereby underlining its pivotal role in clinical scenarios.

To ascertain the clinical utility of this application, we implemented it at the First Medical Center and Nanjing Drum Tower Hospital. From June to October 2023, we prospectively amassed data from 754 patients who underwent cardiac procedures. Throughout their hospitalization, medical professionals diligently utilized this tool, updating patient details in real-time and forecasting imminent CSA-AKI episodes(see Figure 6b). In this period, there were 129 documented AKI events.

Impressively, our model preemptively identified 121 (93.8%) of these episodes for all prediction windows, albeit at the cost of 244 false positives. When predictions were spot-on, the REACT system granted clinicians an average lead time of 16 hours for intervention.

## Discussion

Here we demonstrate the practical feasibility of causal deep learning models for early detection of CSA-AKI within the next 48 hours, which can be used to improve perioperative management. Early detection of CSA-AKI is challenging due to the inherently ambitious intricacies of physiological mechanisms^35^, the insensitivity of biomarkers^36,37^, and the rigorous application requirements of predictive models^38,39^. This is despite the recent shift towards novel biomarkers that promise enhanced early detection capabilities for CSA-AKI. However, these methods are not ready for clinical routine use due to their modest discriminative capabilities, invasive nature, and the need for specific kits or expensive equipment^36,40^.

To address these challenges, in this study, we develop and validate REACT, a temporal causal deep learning model trained on simulated RCTs for real-time prediction of full-stage CSA-AKI. Developed from a novel two-phase algorithmic framework, REACT not only achieves superior performance, but also ensures lightweight variable inputs during its application by capturing stable underlying causal structures. The final model, incorporating six causal variables (age, serum creatinine, urea nitrogen, uric acid, lactate dehydrogenase, and creatine kinase enzyme), demonstrated superior prediction accuracy in both training and three independent validation cohorts. Compared to guideline-recommended pathways, REACT detected CSA-AKI on average 16.35 hours earlier in external tests, 14.65 hours earlier in internal validation.

Renal replacement, as the only effective treatment for CSA-AKI, is generally reserved for the most severe patients. In the absence of additional proven effective treatments, management of CSA-AKI has focused on early detection and the implementation of preventive strategies. However, observational data from 12 hospitals confirmed that all recommended AKI prevention strategies were applied in less than 10% of patients in routine clinical practice^41,42^. This low adherence may be attributed to the lack of a precise intervention “alert” in the guidelines, making the implementation of preventive strategies for CSA-AKI challenging. The limited ability of humans to process complex information and its challenge for clinicians to discern subtle changes in patients at an early stage makes it difficult to discern and capture subtle changes in patients from a large array of patient information. In addition, the inherent complexity of a patient undergoing cardiac surgery, combined with the inherently traumatic aspect of such procedures, can occasionally make a patient’s progress abrupt and sharp. However, static risk scores or models based on baselines fail to capture the real-time progress of patient precipitating shifts during hospitalization^10,43^. Tools focus on predicting the binary event “yes or no” to guide the early intervention windows is vague and bewildering. Our model addresses this issue with a more detailed and unambiguous time window, capturing patient heterogeneity with personalized scores, and more targeted and timely preventive interventions.

In addition, the pathophysiology of CSA-AKI is complex, encompassing micromanipulation, toxins, metabolic, hemodynamic and inflammatory factors, ischemia-reperfusion injury, and oxidative stress^44,45^. These injury mechanisms may not only be interconnected, but also exhibit synergistic behavior characterized by dynamic inter-correlation. Thus, the prediction and interpretation of CSA-AKI involves numerous complex aspects and parameters, making it a high-dimensional, decision-making problem. Previous studies have paid little attention to model interpretation and are mostly correlation-based, providing limited insight into novel markers and pathways beyond prior knowledge^46–48^. For simpler medical scenarios, machine learning and neural networks may be sufficiently effective. However, when these algorithms are applied to dynamic settings with complex physiological mechanisms, methods based on correlational reasoning often risk conflating correlation with causation, significantly limiting the accuracy and stability of the predictions^14^. To address the above challenges, we propose a two-phase algorithmic framework that combines the universal approximation capability of neural networks with a causal discovery module to eliminate confounders. These two-phase algorithmic synergistically evolve during the training process, the first phase leverages the model’s substantial fitting capabilities to construct resilient and robust causal graphs. While the second phase mimics RCTs, sequentially conducts simulated interventions on all variables, and steer the prediction model to evaluate their impact on outcomes to progressively isolate the most contributory variables. Our results demonstrate the superiority of combining causal discovery and deep neural networks over associative reasoning alone in complex, temporal, real-world tasks.

Although some algorithms have been developed for early prediction AKI in intensive care unit (ICU) settings; however, their efficacy is greatly dependent on the quality of the data fed into the model and they often struggle to be applied. For example, Tomašev and colleagues^38^, the most promising in AKI prediction, their developed model that require inputs from hundreds to thousands of features, including the historical medical records of the patient. In practical applications, the absence of a single feature or changes in data structure can potentially lead to a decline in the overall performance of the model. Furthermore, due to privacy and ethical considerations in hospitals, most clinically developed predictive tools are typically used locally, necessitating substantial computational power during model running. On an even more fundamental level, not all predictors are available in all datasets, which limits applications to other institutions. Furthermore, data mismatch between data distributions, typically between training and test sets or development and deployment environments (lab and real-world clinical environments), tends to hurt the generalizability of learned models. As in the case of Tomasev and colleagues, 94% of the participants were male. Subsequent validation on a gender-balanced cohort demonstrated a decrease in predictive performance for women, potentially due to data mismatch and interpretation in the training data^18^. This very rationale inspired our shift towards causal deep learning. While employing a comprehensive, multi-scale, and extensive dataset for model training, REACT distills the complex temporal dynamics among variables into six inexpensive, readily available, objective variables as input by analyzing the causal relationships buried in the temporal dimension. This streamlined approach significantly advances the transition from machine learning models to genuine clinical utility. Moreover, the applicability and practicality of our model is validated in small-scale, prospective, multi-center studies.

Nevertheless, our study has limitations. Being a retrospective study, our study is subject to inherent biases. Despite favorable performance in external and temporal validations, further validation in other population studies is necessary to confirm the risk prediction tool’s effectiveness in improving clinical outcomes.

In conclusion, our study presents a pioneering model that integrates deep learning with causal discovery, facilitating continuous and advanced prediction of AKI up to 48 hours ahead of clinically significant window. This model significantly reduces the number of input variables required during its application, shifting the computational burden and the need for complex variable inputs to the training phase. The REACT model, which embodies this causal deep learning approach, demonstrates immense potential for wider applications beyond its current scope. For routine diagnosis and prediction in an ICU setting, intervention times can be substantially advanced, allowing tailored treatment to start as soon as possible. In addition, its low implementation cost enhances its potential to spread across different economic levels in country and regional medical centers.

## Methods

### Ethics statement and general clinical dataset

In this study, we adhered to the TRIPOD (Transparent Reporting of a Multivariable Prediction Model for Individual Prognosis or Diagnosis) guideline and the “Ensuring Fairness in Machine Learning to Advance Health Equity” checklist to report prediction models (see *Supplements A, B*). We utilized the First Medical Center (from 2000 to 2021) and the Third Medical Center of Chinese PLA General Hospital (from 2000 to 2019) cardiovascular surgery cohort for predictive model derivation, we randomly allocated 80% of the patients from these centers to the training set and reserved the remaining 20% for internal validation. Subsequently, we tested the model using data from three independent hospitals (The Sixth Medical Center of Chinese PLA General Hospital, the seventh Medical Center of Chinese PLA General Hospital, and Nanjing Drum Tower Hospital) spanning 2000-2019. The surgical data for all five hospitals were extracted from electronic medical records (Hospital Information System, HIS), an electronic medical system encompassing comprehensive data on procedures, encoded diagnoses, and laboratory values. It is noteworthy that in 2019, the Chinese PLA General Hospital underwent a reorganization, merging eight previously independent hospitals. Consequently, the patients from the Third, Sixth, and Seventh Centers included in this study were admitted before this consolidation, ensuring that the patient populations were distinct and retained their unique characteristics. All consecutive eligible patients were recruited under a waiver of informed consent. This study actively consented eligible patients and was approved by the institutional review board of the Chinese PLA general hospital (Identifier: S2021-305-01). This study was also approved by the Ethical Review Committee of Nanjing Drum Tower Hospital (Identifier: S2020-281-01).

### Participants and data sources

The study population included adult patients (aged 18 years and older) who underwent coronary artery bypass grafting (CABG), valve surgery, aortic surgery, pericardial surgery, and heart transplantations. We excluded patients who had transcatheter surgery, except for those who received transcatheter aortic valve implantations (Figure 2). We characterized patient comorbidities and procedures using the International Classification of Diseases, Ninth Revision (ICD-9), and the International Classification of Diseases, Tenth Revision (ICD-10) codes. For individuals with multiple admissions, we treated each admission as an independent data point. We excluded patients on long-term dialysis, those requiring preoperative dialysis (up to 6 months before surgery), or those with preoperative serum creatinine values of 4 mg/dL or higher. Furthermore, we excluded patients who developed moderate to severe AKI or initiated dialysis at or before the first postoperative metabolic panel blood draw.

### Outcome measures

The primary endpoint of our study was the occurrence of all-stage AKI, defined according to the modified KDIGO criteria for AKI diagnosis. The criteria specified an increase in serum creatinine by at least 50% within seven days or at least 0.3 mg/dL within 48 hours after cardiac surgery compared to pre-PCI serum creatinine levels. We used the most recent preoperative serum creatinine value as the baseline. The secondary endpoint was AKI requiring dialysis.

### Data preprocessing

In this study, we initially screened participants using 227 surgical codes to account for the potential loss of patients due to incomplete ICD code documentation. We meticulously extracted the names of all surgeries meeting the study criteria and further assessed patients by identifying keywords within surgical names, encompassing a total of 78 items. To refine the study population, we diligently examined billing records during patients” hospital stays, pinpointing 289 surgery-related and consumable charges pertinent to the target group.

To ensure data accuracy, we performed secondary verification of electronic medical record (EMR) text within the database and used Structured Query Language (SQL) to search and construct the database from the Hospital Information System (HIS), with codes reviewed by clinical experts and database engineers. Extracted data were cross-checked against patients” scanned medical records using a random sampling strategy, and search strategies were adjusted based on sampling results. Simultaneously, secondary verification of the data was conducted using relevant epidemiological surveys and high-quality randomized controlled trials (RCTs), and cohort studies. Multiple approaches were employed to process and supplement patients” medical history and diagnoses, detecting and correcting outliers to ensure data accuracy and reliability.

#### Comorbidities

To guarantee data reliability and validity, we integrated ICD-9 and ICD-10 disease codes and utilized natural language processing tools and manual verification to further process patients’ past medical history and address potential omissions of complications. For underreported comorbidities such as hypertension and diabetes, we confirmed diagnoses by reviewing patients” long-term medication prescriptions and referring to standard definitions. (For example, when determining whether a patient has concomitant hypertension, the researchers extracted data from patients using antihypertensive drugs and conducted statistics and analysis on patients who met the diagnostic criteria. As heart failure patients are routinely treated with angiotensin-converting enzyme inhibitors (ACEI) or angiotensin II receptor blockers (ARB) to improve prognosis, this study determined whether there were complications of hypertension by retrieving vital sign information during hospitalization and manually search for relevant cases.)

#### Laboratory examinations

To ensure data accuracy and consistency for patients from multiple medical centers over a 20-year period, we meticulously merged and standardized test items for the same laboratory results according to the sample source, measurement unit, and reference range. We compared and converted different test names, units, and test kits to ensure consistency in meaning.

#### Detection and correction of outliers

We conducted outlier detection and correction to ensure data accuracy, identifying physician input errors based on medical common sense and implementing corrective measures. We also constructed related new variables to further inspect and correct the data. Additionally, we employed machine learning outlier detection methods, such as the multivariate Gaussian distribution, to process and examine structured data. These techniques enabled a more detailed identification of potential anomalies in the data, and combined with other patient information and clinical experience, allowed for further analysis of suspicious data, improving data quality and credibility.

#### Sequential data transformation and construction

To enable dynamic real-time early warning for CSA-AKI, preprocessed data were transformed into temporally structured sequences, enhancing the applicability of deep learning algorithms. Patient hospitalization data, including vital signs, and laboratory test results, were chronologically arranged to create time-series data. We do not use any kind of imputation since the observed data points are sparse. Instead, we indicate the missing points with additional “presence” features. If an indicator has been measured several times, the average measured value of this indicator in the current time window is calculated as the characteristic value of the current time window. Unreasonable time intervals were identified and corrected. It is noteworthy that the measurement time information for the indicators may be stored in two fields: sample reception time and sample report time. In cases where both are available, researchers prioritize the sample reception time as the temporal information for the indicator. For potential manual input errors, such as incorrect months or years, or implausible intervals between measurements and reporting, researchers identified and corrected unreasonable time gaps.

#### AKI labeling

In our study, the AKI categorization was derived from the KDIGO guidelines. This categorization was used to define the present status for each time interval. The subsequent 48-hour statuses were then utilized as labels. Given the progressive nature of AKI, it is noteworthy that some patients might directly present with AKI stage II or III. In such instances, the initial AKI detection time is considered as the onset of AKI stage I. Similarly, if a patient is observed to be at AKI stage III, the earliest AKI detection time is taken as the onset for both AKI stage I and stage II. Additionally, auxiliary labels were incorporated for multi-task learning purposes.

### Causal deep learning

#### Causality in deep learning

Causality is an emerging aspect in deep learning. It mitigates the potential for deep learning models to make data-driven errors by considering causal relationships concealed within complex distributions, and moreover, ensures stability and interpretability across different environments. As an example, we can consider training a neural network to classify cats from dogs, the model may learn to identify a dog by examining whether it is on grass, as many pictures of dogs depict them running on grass. However, this prediction becomes unstable since there may also be pictures of dogs without grass^16^. This issue extends to medical outcome prediction as well, such as predicting CSA-AKI. Relying solely on a patient’s age for prediction is unstable, as it may result in incorrect predictions when young patients develop CSA-AKI due to other reasons. Conversely, if predictions are made based on the risk factors that actually cause the outcomes^49^, e.g., predicting dogs by looking at its ears and noses, predicting CSA-AKI with actual reasons that cause CSA-AKI, the deep learning model becomes more generalizable across different environments.

#### Structural causal model (SCM)

Causality in time-series can be represented as structural causal model (SCM)^50^ by taking into account spatio-temporal structural dependency. We denote a uniformly sampled time-series as 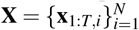, where **x**_*t*_ is the sample vector at time t and consists of N variables{ *x*_*t,i*_ }, with *t* ∈ {1, …, *T*} and *i*∈{ 1, …, *N*}. Then the structural causal model (SCM) for time-series^24^ is

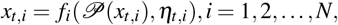

where *f*_*i*_ is a (potentially) nonlinear function that represents spatio-temporal structural dependency, *η*_*t,i*_ denotes latent variables (which can also be considered as noise), and 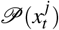 denotes the causal parents of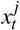. The task for causal discovery (also for our causal deep learning approach) is to identify those causal parents for each variable. In *Supplements S*, we briefly discuss existing approaches for causal discovery.

#### Nonlinear Granger causality

Granger causality is a major class of approaches for causal discovery. We denote by 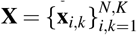 our input dataset (for simplicity we treat all input data as time series here; for static variables such as age or sex, we assume no causal effects from CSA-AKI to these variables, which also satisfies the assumptions of Granger causality), in which **x**_*i,k*_ represents the *i*th time series for patient *k*, with *k* ∈ {1, …, *K* }and *i*∈ {1, …, *N*}, *K* being the total patient number and *N* the time series number for a patient. In this paper, we adopt the representation of existing works^23,25^, and the predicted probability for label *Y*_*k*_ is

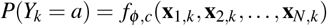

where *i* = 1, 2, …, *N, c* = 1, 2, …, *M, M* is the number of classes. In this paper, we focus on dealing with discovery of causal relationships from **X** to **Y**. For a dynamic system, time-series *i* Granger causes future outcomes **Y** when the past values of time-series *x*_*i*_ **aid in** the prediction of the future status of label **Y**. The standard Granger causality is defined for linear relation scenarios, but recently extended to nonlinear relations^23,25,51^:

##### Definition 1.

Time-series *i* Granger cause outcome **Y** if and only if there exists *c* and 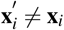,

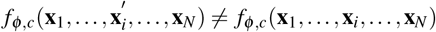

i.e., the past data of time-series *i* influences the prediction of **Y**.

The nonlinear Granger causality is highly compatible with neural networks (NN). Considering the universal approximation ability of NN^28^, it is possible to fit a causal relationship function with component-wise MLPs or RNNs/LSTMs. Moreover, by imposing a sparsity regularizer onto the weights of network connections, as mentioned by^23,52^, NNs can learn the causal relationships from all *N* variables to prediction label. Although Granger causality is not necessarily true (Pearl) causality^53^, it is proved to support causal conclusions when assuming no instantaneous effects and no latent confounder^54^. And recently Granger causality has been applied to a variety of applications because it discovers the causal parents of the interested targets^55^.

#### The two-phase framework of REACT

In this paper, we propose an approach to a) discover variables with causal influences on the interested outcomes and, b) predict the outcomes with the discovered causal variables. These two phases are performed jointly and boosted mutually. In the following section we analyze why causal discovery helps learning with explainability and generalizability. We refer to our algorithm as REACT. To demonstrate our algorithm, we first denote the Causal Probability Graph (CPG) as *G 𝒢*= ⟨*X*, **m**⟩ where the element *m*_*i*_ ∈**m** represents the probability of causal influence from **x**_*i*_ to *Y*, i.e., *m*_*i*_ = *p*(**x**_*i*_ *→Y*). During training, we alternatively learn the prediction model and CPG matrix, which are respectively implemented by *Prediction model training phase* and *Causality learning phase*. We show the implementation details of REACT in *Supplements T*.

#### Prediction model training phase

The proposed *Prediction model training phase* is designed to predict medical outcomes with a neural network *f*_*φ*_, whose structure is shown in Extended Figure 1. The inputs to the neural network include all the historical data points **x**_*i*_ (*i* = 1, 2, …, *N*), and the discovered CPG. During training we sample the causal graph with Bernoulli distribution, in a similar manner to^25,56^’s work, and the predicted probability *p*_*i,c*_ is the output of the neural network *f*_*φ,c*_

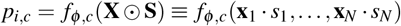

where *s*_*i*_ ∼ Ber(1− *m*_*i*_). **S** is sampled for each training sample in a mini-batch. In this phase, input mask **S** is used as a Dropout-like regularizer^34^ to improve the robustness of the neural network. During training, we update the network parameters *φ* by minimizing the Focal Loss^57^ function ℒ_pred_.

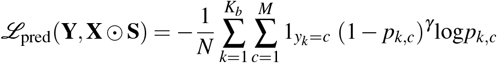

where *K*_*b*_ is the sample number in a mini-batch, *M* is the number of classes, and 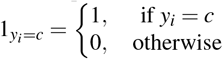. Since the class label for CSA-AKI is highly imbalanced, Focal Loss can effectively address the problem by assigning higher weights to difficult and misclassified examples. The output probability for class *c* from the softmax layer is *p*_*i,c*_ = *f*_*φ,c*_(**X** ⊙ **S**).

#### Causality learning phase

After the *Prediction model training phase*, we proceed to learn CPG in the *Causal Discovery Phase*, to determine the causal probability *m*_*i*_ = *p*(**x**_*i*_ → *Y*), we model this likelihood with *m*_*i*_ = *σ* (*θ*_*i*_) where *σ* (·) denotes the sigmoid function 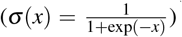 and *θ* is the learned parameter set. Moreover, since the input data **X** always temporally precedes the outcome *Y*, it is unnecessary to learn the edge direction in CPG. Since *s*_*i*_∼ Ber(1− *m*_*i*_) are discrete variables and cannot be directly optimized, we leverage the Gumbel-Softmax technique^58^. We optimize the graph parameters theta by minimizing the following objective

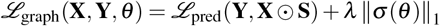

where *ℒ*_pred_ is the Focal Loss penalizing prediction error defined above and ∥·∥_1_ being the *ℒ*_1_ regularizer to enforce sparse connections on the learned CPG. If *θ*_*i*_ are penalized to − ∞ (and *m*_*i*_ → 0), then we deduce that time-series *i* does not Granger cause *Y*. We further prove in the *Supplements R* that under certain assumptions, the discovered causal vector will converge to the true Granger causal relationships.

#### Simulated interventions

In the *Prediction model training phase*, input mask **S** is sampled with Bernoulli distribution and serves as a regularizer. While in this phase, the sampling is used to mimic randomized controlled trials (RCTs) by conducting simulated interventions on each of the variables. Specifically, we intervene on the model by randomly including and excluding a variable and evaluating the causal effects on the outcomes. Supplement R demonstrates that for variables that are actually cause the outcome, the algorithm tends to keep them. Conversely, for spurious variables, the algorithm will gradually eliminate them. As a result, causal variables are identified.

#### Selection of parameter *λ*

Causal threshold parameter *λ*, also determines the number of included variables. A smaller *λ* incorporates more variables with diminished causal effects to the outcome, potentially capturing inter-variable dynamics with spurious connections. Consequently, an excessively small *λ* curtails the model’s transferability, while an overly large *λ* might compromise prediction accuracy. Thus, the selection of *λ* delineates a balance between model capacity and transferability, shown in Figure 5. When *λ* increases from 5× 10^−5^ to 5 × 10^−3^ and the number of variables decreases from 59 to 6, the external testing performance increases drastically while the internal validation performance changes only slightly, as shown in Figure 5 and *Supplements H - K*. This proves that our causal deep learning approach does possess the ability to largely increase transferability while maintaining model capacity with appropriately chosen *λ*. To further reveal the underlying mechanism, local and global feature importances are analyzed by calculating causal effect (Figure 5). We observe that by setting a larger *λ* (i.e., 5 × 10^−3^), fewer causally-important variables are included, which helps increase tranferability since these variables are the most stable predictors. However, *λ* should not be too large because it hampers prediction accuracy too much (*λ* = 5× 10^−2^ for 1 variable model in Figure 5).

#### Generalizability of our model

The sampling strategy (*s*_*i*_ ∼Ber(1 − *m*_*i*_)) in our REACT, not only enables the incorporation of CPG into the neural network, but also prevents the neural network itself from overfitting since it also serves as a regularization strategy like Dropout^34^. As a result, our model enhances the performance and generalizability in the following two aspects, i) Select the robust causal features in *Causality learning phase*. This is empirically validated in Figure 5, which demonstrates that with only six causal variables, the generalizability increases. ii) Prevent the neural network from overfitting in the *Prediction model training phase*. Experimentally, we show in Figure 5 that when training another neural network with only causal features (without our causal deep learning strategy), the performance is still lower than ours. This demonstrates that REACT not only identifies causal variables to enhance generalizability, but also avoids overfitting during the training procedure.

#### Importance analysis via causal inference

With the discovered causal graph, we analyze the importance of each causal variable by calculating average causal effects and counterfactuals. Since all dynamic variables temporally exceed outcomes *Y*_*k*_, there are no edge from *Y*_*k*_ to **X** in the graph model. Moreover, if we assume there are no unobserved confounders, then all backdoor paths from **X** to *Y*_*k*_ is blocked by conditioning on variables excluding *x*_*i*_^55^, and the direct effect is identified as

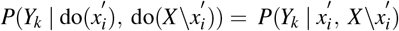

where 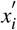 is the reference value of *x*_*i*_,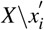 is the set of variables excluding *x*_*i*_′. In Figure 6c and *Supplements U*, we specifically analyze the counterfactual prediction 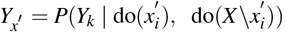. Then individual importance (IIM) is defined as controlled direct causal effect^59^

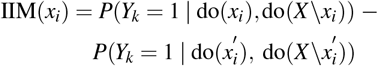

which is used in Figure 5b to demonstrate the importance of a variable for a certain patient. To calculate the average importance for all patient, we have

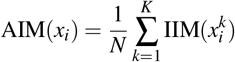

where k denotes the sample index, N denotes the total sample number. Note that our importance analysis approach has limitations. We are based on the assumption that there is no unobserved confounder, which may be too strong in practice. If this assumption does not hold, our causal inference analysis may be biased, and the calculated result is only occlusion analysis of the deep learning model^60^.

### Model capabilities and clinical application

The REACT can generate AKI occurrence probability curves for both patients without AKI and those with existing AKI within the next 48 hours from the current time point. Concurrently, it outputs probability values for AKI occurrence or progression within future 6, 12, 24, and 48-hour time frames. This methodology provides personalized predictions, enabling physicians to efficiently monitor patient progression and determine optimal intervention timings.

Our joint prediction model training and causality learning enable the identification of important features with significant causal influences. This enables us to develop a lightweight model by ignoring most features without significant causal influences (by setting an appropriate *λ*). This lightweight version not only reduces data collection burden, hardware requirements, and deployment complexity but also expedites the model training and inference process, promoting widespread adoption in diverse medical institutions.

To improve the model’s transferability across medical institutions with varying data collection completeness, the lightweight version utilizing a limited set of high impact, easily collected variables was developed. This streamlined model reduces data collection burden, hardware demands, and deployment complexity while accelerating training and inference, promoting widespread adoption in diverse healthcare settings.

### Model evaluation metrics and comparisons

The Area Under the Receiver Operating Characteristic Curve (AU-ROC) was employed to gauge the model’s predictive accuracy, a well-established metric for classification models, representing the area beneath the Receiver Operating Characteristic (ROC) curve. Higher AUROC values indicate superior classifier performance and efficacy. In addition, the Area Under the Precision-Recall Curve (AUPRC) is another commonly used metric for evaluating classification models. The AUPRC quantifies the overall performance of the classifier by calculating the area beneath the Precision-Recall Curve. Higher AUPRC values indicate superior classifier performance, especially in scenarios where the identification of positive instances is of greater importance or when dealing with imbalanced datasets.

Discrimination and calibration, essential performance indicators in model evaluation, measure classification performance and prediction accuracy. Discrimination evaluates the model’s effectiveness in distinguishing between different sample classes, assessed through classification accuracy, the ROC curve, AUROC values, and the Precision-Recall Curve (PRC). High discrimination models demonstrate robust classification capabilities, accurately identifying positive instances while minimizing false positives. Calibration measures the consistency between model predictions and actual outcomes, typically assessed through prediction error, residual analysis, and log-likelihood indicators. High calibration models yield results closely aligned with actual outcomes.

## Data availability

Data underpinning this study are under restricted access and are not freely available as they contain patients’ data, and specific clearance from the ethics committee is required in each center. Data can, however, be made available upon reasonable request. Specific conditions and restrictions of access to the datasets are to be discussed directly with the main investigators in each center.

## Code Availability

The code for REACT is available at https://github.com/jarrycyx/UNN/tree/main/REACT, which includes implementation for data preprocessing, causal discovery, training, validation, and testing of CSA-AKI prediction.

## Author Contributions

Kunlun He, Qionghai Dai and Jinli Suo conceptualized and supervised the study. Qin Zhong, Yuxiao Cheng, and Zongren Li were responsible for the study design, execution of all project components, and manuscript preparation. Dongjin Wang provided a dataset from Nanjing Drum Tower Hospital for external tests and offered guidance on data curation as well as in writing the clinical discussions. Chongyou Rao and Yi Jiang contributed expert clinical opinions in case selection, data collection, and manuscript review. They also assisted in manual data validation and the compilation of the final dataset. Lianglong Li, and Ziqian Wang participated in the development, debugging, and optimization of the model. Pei Li conducted the final formatting and grammatical checks.Pan Liu took charge of developing and maintaining the website. Yawei Zhao participated in organizing the prospective validation of the study at two centers, which implies involvement in coordinating and managing parts of the research process.

## Acknowledgements

None of this material has been published or is under consideration for publication elsewhere. We declare no competing financial interests. This research was supported by the National Key Research and Development Program of China (2021ZD0140406) and project of the Medical Engineering Laboratory of Chinese PLA General Hospital [2022SYSZZKY20, 2022SYSZZKY11]. The funders had no role in considering the study design or in the collection, analysis, interpretation of data, writing of the report, or decision to submit the article for publication.

## Ethics declarations

The authors declare no competing interests.

**Extended Figure 1.**
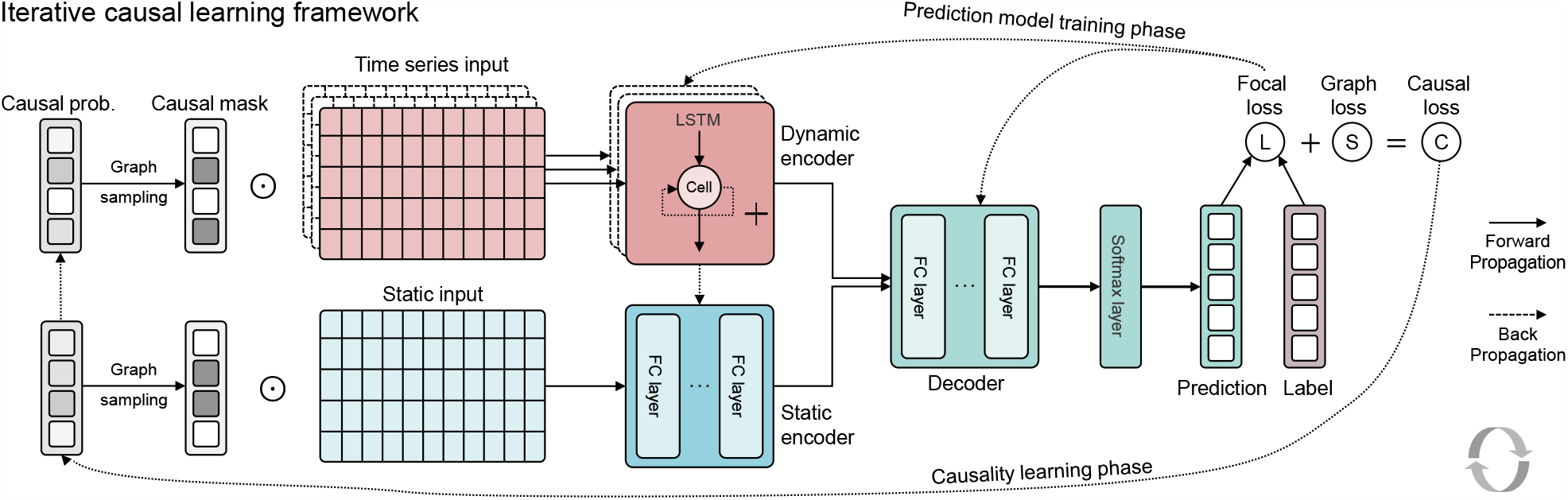
Illustration of iterative causal deep learning process. Iterative causal deep learning. Our model consists of two separate phases with EM-like training strategy. The Prediction model training phase predicts risks for AKI at each time-points given time-series and static input. The Causality learning phase learns causal probability graphs with a fixed prediction model. The causal deep learning eventually generates highly-accurate prediction models by only taking into account causal features, which then boosts the external (or out-of-distribution) testing performance.

**Extended Figure 2.**
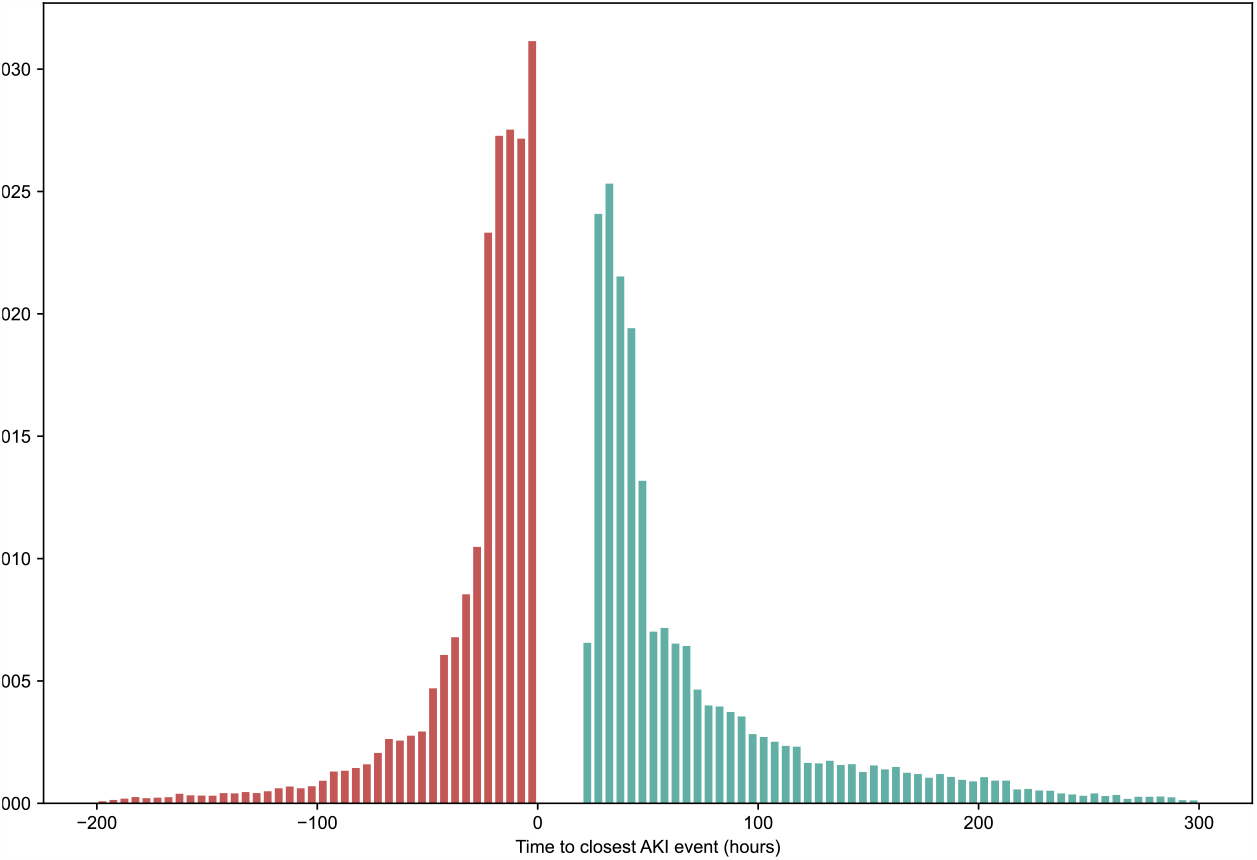
Illustration of false-positives by analyzing the assessment time to closest AKI event. We show the results by predicting all AKI within 24 hours, thresholded at 33% precision. We show the histogram of actual AKI events before or after the time of assessment, excluding those within 0 to 24 hours after time of assessment (true positives). We observe 34.4% of false positives are attributed to positive prediction after AKI, 8.2% are attributed to 24-48 hours after time of assessment. Only about half (50.7%) are actual false positives and no AKI occurs at any point for this patient.

**Extended Figure 3.**
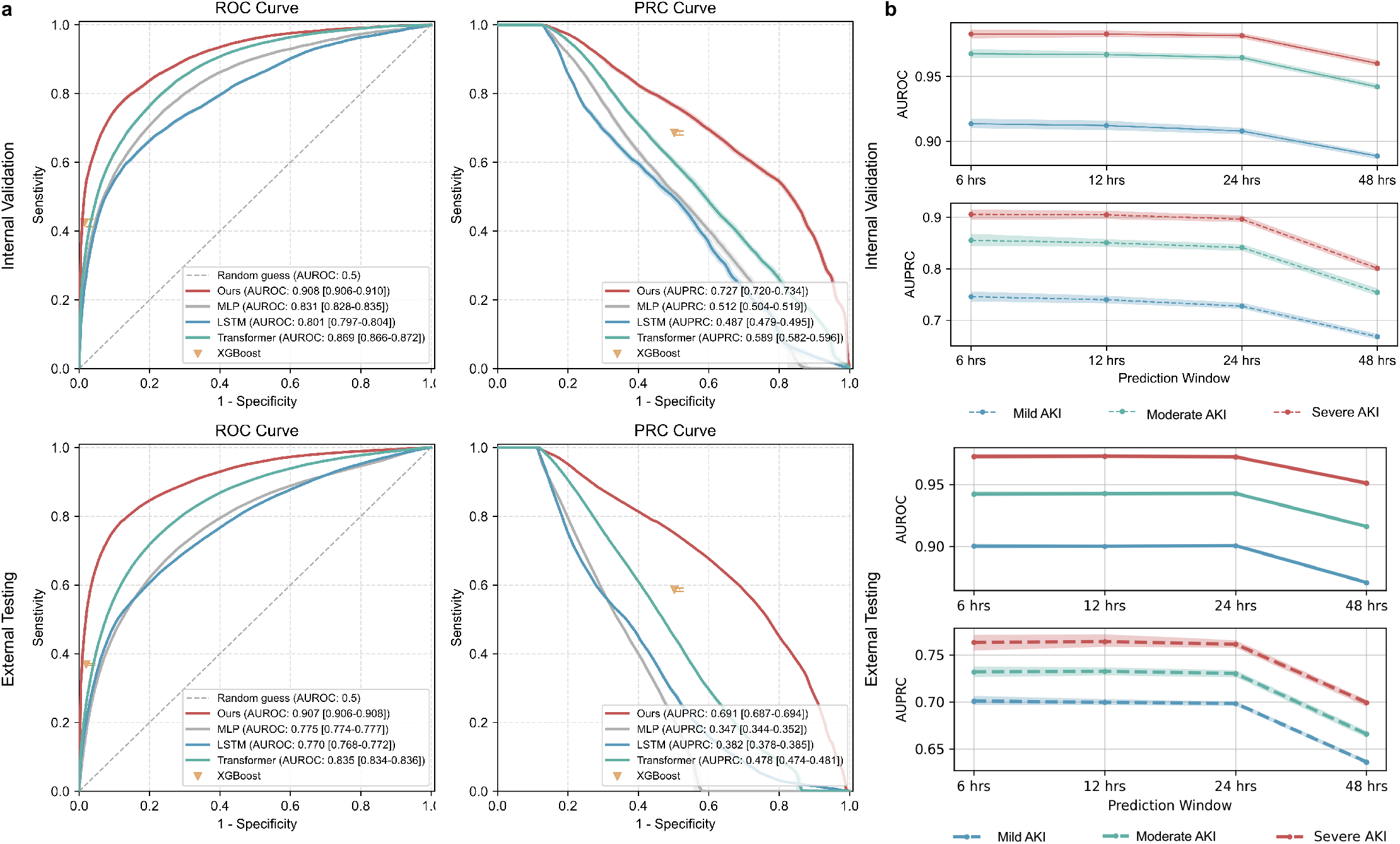
Model performance to validate longitudinal transferability. **a**, Receiver-operating characteristic curves and Precision-recall curves for mild AKI within 24 hours, on internal validation (randomly allocated) and external testing datasets (2018-2021, which is not included in the training and validation datasets), comparing with baseline methods without casual learning (see Methods). **b**, AUROC and AUPRC on other prediction tasks and windows, i.e., 4 kinds of prediction windows (6, 12, 24, 48 hours), each with 3 types of AKI (mild AKI, moderate AKI, and severe AKI).

**Extended Figure 4.**
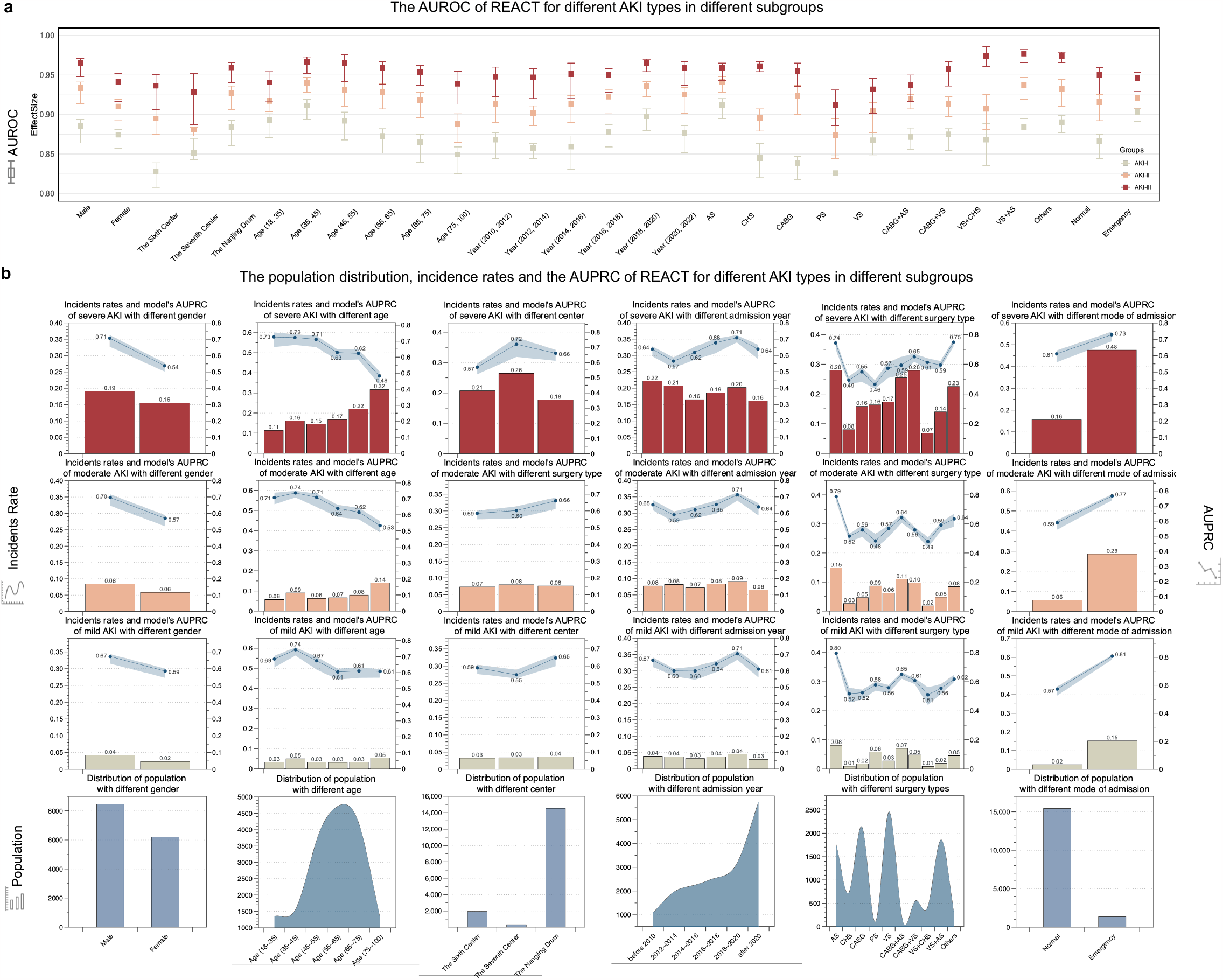
Model performance of different patient cohorts in external testing datasets. **a**, Displays the AUROC predictions of REACT across all subgroups. Different colors signify the model’s performance at various CSA-AKI stages, with the vertical range indicating the model’s prediction performance variability within 48 hours of the event occurrence. The corresponding abbreviations in the operation type subgroups are as follows: AS, Aortic surgery; CHS, congenital heart disease corrective surgery; CABG, coronary artery bypass grafting; PC, pericardiectomy; VA, valve surgery. **b**, Illustrates the AUPRC predictions of REACT in different subgroups and provides detailed information about the population and incidence rate within each subgroup. Transverse subgraphs 1-5 specifically depict the model’s performance across varying surgical types, medical centers, age groups, admission years and genders. Longitudinal subgraphs 1-3 use bar graphs to show the incidence rate of CSA-AKI stage III-I among patients of different subgroups (as denoted on the left y-axis) and use plots to show the corresponding CSA-AKI prediction AUPRC with the shadow indicating performance variability within 48 hours of the event(as denoted on the right y-axis); In Longitudinal subgraph 4, the total number of patients in each subgroup is represented.

**Extended Figure 5.**
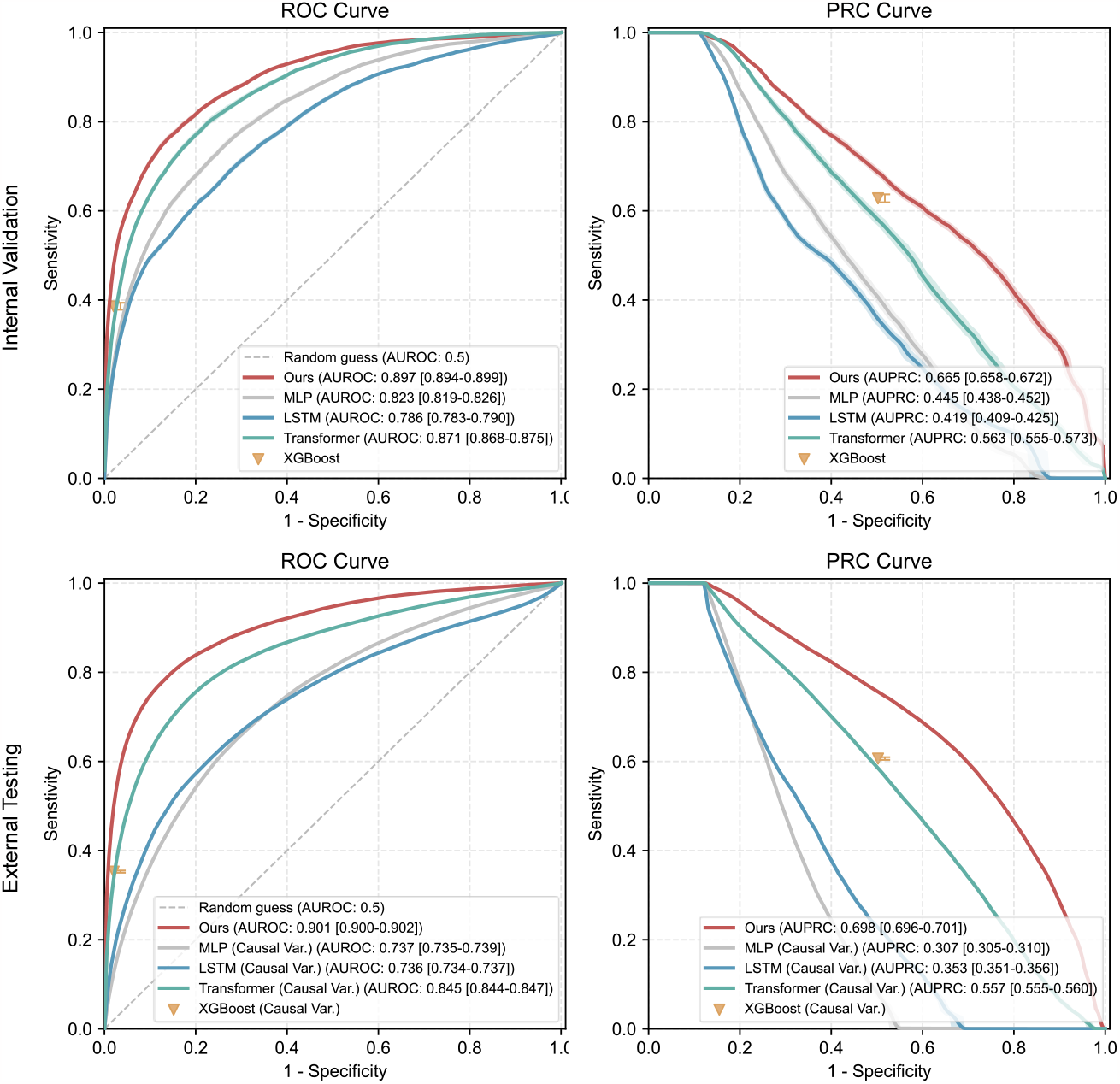
Model performance comparing with baselines using only causal variables. Receiver-operating characteristic curves and Precision-recall curves of different methods for mild AKI within 24 hours, on internal validation and external testing datasets.

**Extended Figure 6.**
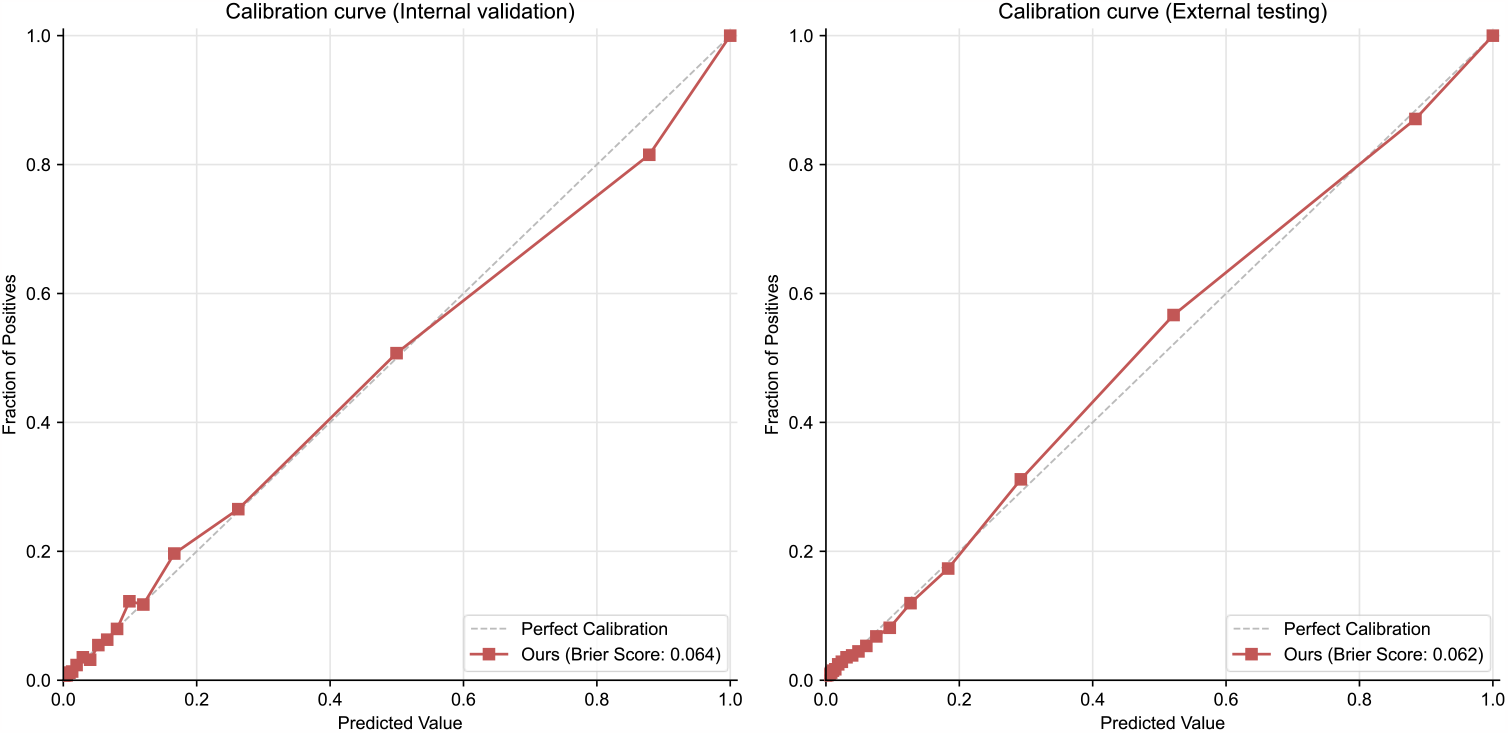
Model calibration. Our model is well calibrated, here show calibration curves and Brier Score on internal validation and external testing set.

## Notes

### Competing Interest Statement

The authors have declared no competing interest.

### Author Declarations

Institutional review board of the Chinese PLA general hospital gave ethical approval for this work Ethical Review Committee of Nanjing Drum Tower Hospital gave ethical approval for this work

