## Supplementary Information for "Ultra-efficient causal deep learning for Dynamic CSA-AKI Detection Using Minimal Variables"

The purpose of this supplementary information is to offer additional insights that substantiate the claims made in the paper titled " Ultra-efficient Causal Learning for Dynamic CSA-AKI Detection Using Minimal Variables". The authors anticipate that by providing this supplementary data and corresponding discussions, the strength and reproducibility of the findings presented in the main letter will be enhanced. In conjunction with the Extended Data, the following supplementary materials are introduced:

- Supplements A and B display the normative guidelines followed in the design, development, and validation phases of this research, as well as their corresponding placements in the main text.
- Supplement C offers a detailed view of the demographic information of all patients included in the model's training, validation, and testing stages.
- Supplements D-E display the variables included in REACT training and show the definitions of certain clinical variables.
- Supplements F-G shows the performance of REACT and comparisons with current mainstream algorithms and elaborate on the differences between models using all variables and those only inputting causal variables.
- Supplements H-K present the predictive performance of REACT across various sub-populations in both internal validation and external test datasets.
- Supplements L present the number of input variables and the model performance of REACT with different λ.
- Supplements M-P delineate the sensitivity and specificity of the prediction system when different thresholds are selected.
- Supplement Q compares outcomes between patients observed with AKI stage I occurrences and those who were not observed.
- Supplements R-T provide the underlying theorems and proofs relied upon by the algorithm, intricate algorithmic details, parameter information, and related work.
- Supplement U provide some other examples with counterfactual explanations.

### *A. TRIPOD checklist (Prediction Model Development and Validation)*

| Section/Topic | | | Checklist Item | Page |
|---|---|---|---|---|
| **Title and abstract** | | | | |
| Title | 1 | D;V | Identify the study as developing and/or validating a multivariable prediction model, the target population, and the outcome to be predicted. | Page 1 |
| Abstract | 2 | D;V | Provide a summary of objectives, study design, setting, participants, sample size, predictors, outcome, statistical analysis, results, and conclusions. | Page 1 |
| **Introduction** | | | | |
| Background and objectives | 3a | D;V | Explain the medical context (including whether diagnostic or prognostic) and rationale for developing or validating the multivariable prediction model, including references to existing models. | Line 39~85 |
| | 3b | D;V | Specify the objectives, including whether the study describes the development or validation of the model or both. | Line 86~101 |
| **Methods** | | | | |
| Source of data | 4a | D;V | Describe the study design or source of data (e.g., randomized trial, cohort, or registry data), separately for the development and validation data sets, if applicable. | Line 86~122, 593~609 |
| | 4b | D;V | Specify the key study dates, including start of accrual; end of accrual; and, if applicable, end of follow-up. | Line 103~122 |
| Participants | 5a | D;V | Specify key elements of the study setting (e.g., primary care, secondary care, general population) including number and location of centers. | Line 103~122 |
| | 5b | D;V | Describe eligibility criteria for participants. | Line 593~609, Fig 2 |
| | 5c | D;V | Give details of treatments received, if relevant. | No relevant |
| Outcome | 6a | D;V | Clearly define the outcome that is predicted by the prediction model, including how and when assessed. | Line 86~101, 186~191,799~816 |
| | 6b | D;V | Report any actions to blind assessment of the outcome to be predicted. | Line 568~577 |
| Predictors | 7a | D;V | Clearly define all predictors used in developing or validating the multivariable prediction model, including how and when they were measured. | Line 828~849, 343~372, Fig 5 |

| | | | | |
|---|---|---|---|---|
| | 7b | D;V | Report any actions to blind assessment of predictors for the outcome and other predictors. | Line 828~849, 343~372 |
| Sample size | 8 | D;V | Explain how the study size was arrived at. | Line 563~592 |
| Missing data | 9 | D;V | Describe how missing data were handled (e.g., complete-case analysis, single imputation, multiple imputation) with details of any imputation method. | Line 618~674 |
| Statistical analysis methods | 10a | D | Describe how predictors were handled in the analyses. | Line 767~816 |
| | 10b | D | Specify type of model, all model-building procedures (including any predictor selection), and method for internal validation. | Line 157~191, 767~816 |
| | 10c | V | For validation, describe how the predictions were calculated. | Line 879~901 |
| | 10d | D;V | Specify all measures used to assess model performance and, if relevant, to compare multiple models. | Line 902~926 |
| | 10e | V | Describe any model updating (e.g., recalibration) arising from the validation, if done. | No relevant |
| Risk groups | 11 | D;V | Provide details on how risk groups were created, if done. | No relevant |
| Development vs. validation | 12 | V | For validation, identify any differences from the development data in setting, eligibility criteria, outcome, and predictors. | Line 767~864, 251~273, 395~421 |
| **Results** | | | | |
| Participants | 13a | D;V | Describe the flow of participants through the study, including the number of participants with and without the outcome and, if applicable, a summary of the follow-up time. A diagram may be helpful. | Line 103~142, Fig 2 |
| | 13b | D;V | Describe the characteristics of the participants (basic demographics, clinical features, available predictors), including the number of participants with missing data for predictors and outcome. | Line 103~142, Sup C |
| | 13c | V | For validation, show a comparison with the development data of the distribution of important variables (demographics, predictors and outcome). | Line 103~142, Fig 2, Sup C |
| Model development | 14a | D | Specify the number of participants and outcome events in each analysis. | Line 286~293, 410~421, Fig 5 |
| | 14b | D | If done, report the unadjusted association between each candidate predictor and outcome. | Line 343~394 |
| Model specification | 15a | D | Present the full prediction model to allow predictions for individuals (i.e., all regression coefficients, and model intercept or baseline survival at a given time point). | Line 251~273 |
| | 15b | D | Explain how to the use the prediction model. | Line 251~273 |

| | | | | |
|---|---|---|---|---|
| Model performance | 16 | D;V | Report performance measures (with CIs) for the prediction model. | Line 192~393 |
| Model-updating | 17 | V | If done, report the results from any model updating (i.e., model specification, model performance). | No relevant |
| **Discussion** | | | | |
| Limitations | 18 | D;V | Discuss any limitations of the study (such as nonrepresentative sample, few events per predictor, missing data). | Line 541~546 |
| Interpretation | 19a | V | For validation, discuss the results with reference to performance in the development data, and any other validation data. | Line 435~448 |
| | 19b | D;V | Give an overall interpretation of the results, considering objectives, limitations, results from similar studies, and other relevant evidence. | Line 423~448 |
| Implications | 20 | D;V | Discuss the potential clinical use of the model and implications for future research. | Line 541~567 |
| **Other information** | | | | |
| Supplementary information | 21 | D;V | Provide information about the availability of supplementary resources, such as study protocol, Web calculator, and data sets. | Line 395~409, 928~933, Sup A,B |
| Funding | 22 | D;V | Give the source of funding and the role of the funders for the present study. | Line 1156~1161 |

**Items relevant only to the development of a prediction model are denoted by D, items relating solely to a validation of a prediction model are denoted by V, and items relating to both are denoted D;V. We recommend using the TRIPOD Checklist in conjunction with the TRIPOD Explanation and Elaboration document.*

#### *B. “Ensuring Fairness in Machine Learning to Advance Health Equity” checklist [REF]*

| Item | Checklist Item | Page |
|---|---|---|
| **Design** | | |
| 1 | Determine the goal of a machine-learning model and review it with diverse stakeholders, including protected groups. | 62~101 |
| 2 | Ensure that the model is related to the desired patient outcome and can be integrated into clinical workflows. | 251~285，395~421 |
| 3 | Discuss ethical concerns of how the model could be used. | 449~472 |
| 4 | Decide what groups to classify as protected. | No relevant |
| 5 | Study whether the historical data are affected by health care disparities that could lead to label bias. If so, investigate alternative labels. | 221~250 |
| **Data collection** | | |
| 6 | Collect and document training data to build a machine-learning model. | 103~142, 563~592 |
| 7 | Ensure that patients in the protected group can be identified (weighing cohort bias against privacy concerns). Assess whether the protected group is represented adequately in terms of numbers and features. | 221~250 |
| **Training** | | |
| 8 | Train a model taking into account the fairness goals. | 86~101, 185~191 |
| **Evaluation** | | |
| 9 | Measure important metrics and allocation across groups. | 192~220, 221~250 |
| 10 | Compare deployment data with training data to ensure comparability. | 286~313, 410~421 |
| 11 | Assess the usefulness of predictions to clinicians initially without affecting patients. | 251~273 |
| **Launch review** | | |
| 12 | Evaluate whether a model should be launched with all stakeholders, including representatives from the protected group. | No relevant |
| 13 | Monitored deployment | 410~421 |
| 14 | Systematically monitor data and important metrics throughout deployment. Gradually launch and continuously evaluate metrics with automated alerts. Consider a formal clinical trial design to assess patient outcomes. Periodically collect feedback from clinicians and patients. | 410~421 |

### C. *The characteristics of patients of our studies*

| | The Derivation Dataset | | The External Testing Dataset | | |
|---|---|---|---|---|---|
| | The First Medical Centers of the Chinese PLA General Hospital (n=12,685) | The Third Medical Centers of Chinese PLA General Hospital (n=1,828) | The Sixth Medical Center of the Chinese PLA General Hospital (n=2,261) | The Seventh Medical Center of the Chinese PLA General hospital (n=1,570) | The Nanjing Drum Tower Hospital (n=16,982) |
| **Patients' demographics,** *Median, [IQR]* | | | | | |
| Age, years | 58.0 [47.0, 66.0] | 45.0 [29.0, 58.0] | 59.0 [48.0, 66.0] | 56.0 [40.0, 68.0] | 57.0[47.0,67.0] |
| Male, count (average) | 8101 (63.9) | 897 (49.1) | 1426 (63.1) | 955 (60.8) | 9824 (57.8) |
| Hight, cm, | 165 [159.0, 171.0] | 164 [160.5, 170.0] | 168 [160.0, 172.0] | 165 [158.5, 170.5] | 165[160.0,171.0] |
| Weight, kg, | 67.0 [59.0, 75.5] | 58.0 [50.0, 68.0] | 66.0 [58.0, 75.0] | 66.0 [59.0, 74.2] | 65[57.3,75.0] |
| **Commodities,** *count (average)* | | | | | |
| Hypertension | 6798 (53.6) | 888 (48.6) | 1328 (58.7) | 609 (38.8) | 10763 (63.4) |
| Diabetes | 3890 (30.7) | 178 (9.7) | 424 (18.8) | 240 (15.3) | 3826 (22.5) |
| Congestive heart failure | 3970 (31.3) | 594 (32.5) | 465 (20.6) | 288 (18.3) | 8480 (49.9) |
| pulmonary disease | 897 (7.1) | 70 (3.8) | 922 (40.8) | 102 (6.5) | 5437 (32.0) |
| Chronic kidney disease | 398 (3.1) | 36 (2.0) | 54 (2.4) | 17 (1.1) | 999 (5.9) |
| **Type of surgery**, *count (average)* | | | | | |
| CABG alone | 4877 (38.4) | 324 (17.7) | 1147 (50.7) | 690 (43.9) | 2031 (12.0) |
| Valve surgery alone | 4263 (33.6) | 712 (38.9) | 618 (27.3) | 351 (22.4) | 5272 (31.0) |
| CABG and valve surgery | 451 (3.6) | 0 (0.0) | 6 (0.3) | 0 (0.0) | 1046 (6.2) |
| Aortic surgery | 776 (6.1) | 14 (0.8) | 135 (6.0) | 94 (6.0) | 3420 (20.1) |
| Congenital Heart Surgery | 1057 (8.3) | 646 (35.3) | 213 (9.4) | 320 (20.4) | 1756 (10.3) |
| Others | 1261 (9.9) | 132 (7.2) | 142 (6.3) | 115 (7.3) | 3457 (20.4) |
| **Surgery characteristics**, *count (average)* | | | | | |

| | | | | | |
|---|---|---|---|---|---|
| Number of surgeries involving cardiopulmonary bypass | 11167 (88.0) | 1565 (85.6) | 1038 (45.9) | 197 (12.5) | 12717 (71.1) |
| Use of intra-aortic balloon pump | 478 (3.8) | 29 (1.6) | 258 (11.4) | 19 (1.2) | 257 (1.5) |
| Use of ECMO | 22 (0.2) | 2 (0.1) | 19 (0.8) | 0 (0.0) | 166 (1) |
| **Preoperative laboratories,** *Median, [IQR]* | | | | | |
| Serum platelet, 10*9/L | 193.0 [156.0, 234.0] | 199.0 [163.0, 241.0] | 204.0 [166.0, 246.0] | 207.0 [168.5, 256.5] | 176.0[138.0,220.0] |
| Mean Corpusular Hemoglobin Concerntration, g/L | 338.0 [330.0, 346.0] | 333.0 [324.0, 340.0] | 338.0 [330.0, 34 5.0] | 329.0 [322.0, 336.0] | 335.0[327.0,342.0] |
| Serum albumin, g/L | 41.0 [38.5, 43.4] | 41.9 [39.2, 44.7] | 40.3 [37.6, 42.7] | 40.6 [37.8, 43.7] | 39.8[37.5,41.9] |
| Serum potassium, mmol/L | 4.08 [3.85, 4.33] | 3.99 [3.74, 4.25] | 3.92 [3.60, 4.20] | 3.96 [3.76, 4.19] | 3.99[3.75,4.24] |
| blood urea nitrogen, mmol/L | 5.64 [4.60, 7.02] | 5.73 [4.58, 7.12] | 5.60 [4.60, 6.90] | 5.54 [4.40, 6.95] | 6.37[5.1,8.21] |
| Serum creatinine, umol/L | 75.2 [64.1, 88.2] | 63.0 [53.0, 76.0] | 84.6 [73.9, 97.0] | 69.0 [58.0, 81.0] | 67.0[56.0,81.0] |

*IQR, Interquartile range; CABG, coronary artery bypass grafting;*

#### *D. The definition of clinically relevant variables*

Demographics, clinical and main surgery characteristics were retrospectively retrieved from patients' medical history and electronic medical charts at each center for the PRIDE and HIS registry for patients enrolled.

*Hypertension* was defined as an history of high blood pressure diagnosed or treated by a physician, being in treatment with anti-hypertensive drugs or admission blood pressure >140/90 mmHg.

*Diabetes mellitus* was defined as an history of diabetes mellitus diagnosed or treated by a physician, being in treatment with hypoglycaemic drugs or an admission Hb1Ac value >6.5% (48 mmol/mol).

*Heart Failure* was defined as there are symptoms and/or signs of congestion in systemic and/or pulmonary circulation, requiring treatment with diuretics.

*Chronic kidney disease (CKD)* was defined as history of chronic renal insufficiency with an estimated glomerular filtration rate (eGFR) < 60ml/min/1.73 $m^2$ for more than 3 months.

*Respiratory disease* was defined as includes chronic obstructive pulmonary disease, chronic bronchitis and sleep apnea hypopnea syndrome.

*Left ventricular ejection fraction* (LVEF) was assessed at discharge by 2D transthoracic echocardiography and computed according to bidimensional Simpson formula [(left ventricular end diastolic volume – left ventricular end systolic volume)/ left ventricular end diastolic volume)].

#### *E. All variables included in the model training process.*

| | | | | | | | |
|---|---|---|---|---|---|---|---|
| Demographic | Chronic Kidney Disease | Ticagrelor | Hematocrit Measurement | Calcium | Urine White Blood Cells Examination (Microscopy) | Fibrinogen Measurement | Urine Yeast Cells |
| Age | Intra-aortic balloon pump (IABP) | Beta Blocker | Mean Corpuscular Volume (MCV) | Direct Bilirubin | Urine Epithelial Cells Examination (Microscopy) | International Normalized Ratio (INR) | Urine 70% Red Cell Forward Scatter Position |
| Gender | Previous MI (myocardial infarction) | Statin | Platelet Count | Partial pressure of Carbon Dioxide | Urine Specific Gravity Measurement | Total Bile Acids | Mean Platelet Volume Measurement (MPV) |
| Body Height | Previous CABG | Bicarbonate | Mean Corpuscular Hemoglobin (MCH) | Urine Small Round Epithelial Cells | Urine pH Measurement | Lactate Dehydrogenase (LDH) | Plasma D-Dimer Measurement |
| Body Weight | Previous PCI (percutaneous coronary intervention) | Glucose | Mean Corpuscular Hemoglobin Concentration (MCHC) | Inorganic Phosphorus | Urine White Blood Cell Examination | Alkaline Phosphatase (ALP) | Creatine Kinase Isoenzyme (CK-MB) ng/ml |
| Surgery information | Previous Stroke | Oxygen saturation | Troponin T | Creatine Kinase Isoenzyme (CK-MB) U/L | Urine Nitrite Test | Gamma-Glutamyl Transferase (GGT) | Thrombin Time Measurement |
| Surgery type | Left Ventricular Ejection Fraction (LVEF) | Partial pressure of carbon dioxide and oxygen | Sodium | Creatine Kinase (CK) | Urine Protein Qualitative Test | Triglycerides | Red Cell Distribution Width Measurement CV (RDW-CV) |
| Emergency surgery | Renal replacement therapy | pH level | Glucose | Magnesium | Urine Glucose Qualitative Test | Total Cholesterol | Direct Eosinophil Count |
| Surgery duration | Systolic arterial pressure | Neutrophil Percentage | Total Protein | Direct Bilirubin | Urine Ketone Test | High-Density Lipoprotein Cholesterol (HDL-C) | Blood Culture (Aerobic) + Identification + Drug Sensitivity |

| | | | | | | | |
|---|---|---|---|---|---|---|---|
| Cardiopulmonary bypass | mean arterial pressure | Lymphocyte count | Aspartate Aminotransferase (AST) | C-Reactive Protein Measurement (CRP) | Urine Biliverdin Qualitative Test | Low-Density Lipoprotein Cholesterol (LDL-C) | Irregular Antibody Screening |
| Intraoperative blood transfusion | diastolic arterial pressure | Monocyte count | Potassium | Fecal White Blood Cells | Urine Bilirubin Qualitative Test | Occult blood test (stool) | Urine Red Cell Forward Scatter Width |
| Comorbidity | Body temperature | Eosinophil count | Blood Urea Nitrogen (BUN) | Fecal Red Blood Cells | Urine Red Blood Cell Examination | Urine Red Blood Cells | Urine Conductivity |
| Diabetes Mellitus | heart rate | Basophil count | Serum Albumin | Fecal Parasite Ova | Urine Casts Examination (Microscopy) | Urine White Blood Cells | Hepatitis B e Antibody (Luminescence Method) |
| Hypertension | Respiratory frequency | White Blood Cell Count (WBC) | Alanine Transaminase | Hepatitis B Surface Antibody (Luminescence Method) | ABO Blood Group Identification | Urine Epithelial Cells | Hepatitis B Core Antibody (Luminescence Method) |
| Respiratory System | ACE inhibitors/ARBs | Red Blood Cell Count (RBC) | Chloride | Hepatitis B e Antigen | Prothrombin Time Measurement | Urine Crystals | RH Blood Typing (D Antigen) |
| Heart Failure | Clopidogrel | Hemoglobin Measurement | Serum Creatinine | Urine Red Blood Cells Examination (Microscopy) | Prothrombin Activity Measurement | Serum Uric Acid | |

#### *F. The AUROC of REACT and other methods at different time points.*

| | Mild AKI | | | | Moderate AKI | | | | Severe AKI | | | |
|---|---|---|---|---|---|---|---|---|---|---|---|---|
| | 6 hrs | 12 hrs | 24 hrs | 48 hrs | 6 hrs | 12 hrs | 24 hrs | 48 hrs | 6 hrs | 12 hrs | 24 hrs | 48 hrs |
| **REACT** | 0.886 / 0.899 | 0.886 / 0.897 | 0.887 / 0.892 | 0.857 / 0.884 | 0.930 / 0.939 | 0.931 / 0.939 | 0.932 / 0.936 | 0.904 / 0.917 | 0.964 / 0.972 | 0.965 / 0.971 | 0.964 / 0.969 | 0.937 / 0.949 |
| **MLP** | 0.693 / 0.831 | 0.688 / 0.829 | 0.679 / 0.823 | 0.666 / 0.830 | 0.726 / 0.871 | 0.721 / 0.867 | 0.711 / 0.862 | 0.704 / 0.855 | 0.755 / 0.915 | 0.748 / 0.916 | 0.739 / 0.913 | 0.738 / 0.901 |
| **LSTM** | 0.753 / 0.780 | 0.755 / 0.782 | 0.758 / 0.787 | 0.737 / 0.802 | 0.806 / 0.810 | 0.808 / 0.814 | 0.809 / 0.820 | 0.794 / 0.825 | 0.848 / 0.871 | 0.849 / 0.874 | 0.850 / 0.880 | 0.832 / 0.875 |
| **Transformer** | 0.840 / 0.881 | 0.839 / 0.878 | 0.835 / 0.872 | 0.799 / 0.876 | 0.892 / 0.921 | 0.892 / 0.920 | 0.889 / 0.912 | 0.855 / 0.901 | 0.927 / 0.948 | 0.927 / 0.949 | 0.926 / 0.943 | 0.899 / 0.931 |
| **MLP(Causal feat)** | 0.717 / 0.813 | 0.719 / 0.813 | 0.719 / 0.809 | 0.707 / 0.819 | 0.713 / 0.836 | 0.711 / 0.836 | 0.717 / 0.831 | 0.719 / 0.830 | 0.710 / 0.864 | 0.711 / 0.861 | 0.726 / 0.859 | 0.738 / 0.865 |
| **LSTM(Causal feat)** | 0.713 / 0.783 | 0.717 / 0.784 | 0.721 / 0.786 | 0.699 / 0.803 | 0.784 / 0.797 | 0.788 / 0.798 | 0.789 / 0.800 | 0.767 / 0.813 | 0.830 / 0.841 | 0.832 / 0.843 | 0.833 / 0.844 | 0.811 / 0.847 |
| **Transformer (Causal feat)** | 0.851 / 0.877 | 0.854 / 0.877 | 0.851 / 0.870 | 0.816 / 0.872 | 0.899 / 0.921 | 0.901 / 0.922 | 0.898 / 0.916 | 0.865 / 0.903 | 0.930 / 0.947 | 0.931 / 0.949 | 0.929 / 0.943 | 0.900 / 0.928 |

*The notation "/" separates the performance metrics of our model on external test sets (before) from its performance on internal validation sets (behind). The term "(Causal feat)" indicates that the model was constructed using causal variables identified through our methodology.*

### *G. The AUPRC of REACT and other methods at different time points.*

| | Mild AKI | | | | Moderate AKI | | | | Severe AKI | | | |
|---|---|---|---|---|---|---|---|---|---|---|---|---|
| | 6 hrs | 12 hrs | 24 hrs | 48 hrs | 6 hrs | 12 hrs | 24 hrs | 48 hrs | 6 hrs | 12 hrs | 24 hrs | 48 hrs |
| **REACT** | 0.652 / 0.663 | 0.653 / 0.654 | 0.654 / 0.637 | 0.593 / 0.599 | 0.667 / 0.688 | 0.668 / 0.682 | 0.669 / 0.670 | 0.603 / 0.607 | 0.668 / 0.739 | 0.670 / 0.737 | 0.670 / 0.724 | 0.606 / 0.665 |
| **MLP** | 0.280 / 0.475 | 0.278 / 0.464 | 0.266 / 0.445 | 0.259 / 0.434 | 0.213 / 0.424 | 0.212 / 0.408 | 0.200 / 0.387 | 0.195 / 0.355 | 0.155 / 0.402 | 0.153 / 0.385 | 0.143 / 0.368 | 0.143 / 0.347 |
| **LSTM** | 0.387 / 0.428 | 0.388 / 0.424 | 0.385 / 0.419 | 0.367 / 0.425 | 0.347 / 0.381 | 0.348 / 0.379 | 0.343 / 0.375 | 0.328 / 0.367 | 0.308 / 0.387 | 0.311 / 0.391 | 0.309 / 0.396 | 0.292 / 0.388 |
| **Transformer** | 0.564 / 0.592 | 0.562 / 0.583 | 0.553 / 0.564 | 0.516 / 0.545 | 0.579 / 0.584 | 0.577 / 0.574 | 0.566 / 0.553 | 0.527 / 0.501 | 0.590 / 0.607 | 0.590 / 0.604 | 0.579 / 0.590 | 0.541 / 0.540 |
| **MLP(Causal feat)** | 0.272 / 0.417 | 0.273 / 0.413 | 0.272 / 0.393 | 0.264 / 0.389 | 0.162 / 0.324 | 0.163 / 0.315 | 0.165 / 0.292 | 0.163 / 0.277 | 0.086 / 0.250 | 0.088 / 0.242 | 0.090 / 0.226 | 0.092 / 0.230 |
| **LSTM(Causal feat)** | 0.289 / 0.388 | 0.290 / 0.383 | 0.288 / 0.375 | 0.271 / 0.379 | 0.226 / 0.298 | 0.227 / 0.292 | 0.223 / 0.285 | 0.210 / 0.278 | 0.158 / 0.271 | 0.158 / 0.269 | 0.156 / 0.271 | 0.150 / 0.278 |
| **Transformer (Causal feat)** | 0.547 / 0.594 | 0.551 / 0.588 | 0.545 / 0.563 | 0.501 / 0.540 | 0.540 / 0.605 | 0.542 / 0.600 | 0.537 / 0.575 | 0.491 / 0.523 | 0.529 / 0.628 | 0.531 / 0.636 | 0.525 / 0.611 | 0.476 / 0.559 |

*The notation "/" separates the performance metrics of our model on external test sets (before) from its performance on internal validation sets (behind). The term "(Causal feat)" indicates that the model was constructed using causal variables identified through our methodology.*

#### *H. The AUROC of REACT at different time points in different subgroups in internal validation.*

| | | Mild AKI | | | | Moderate AKI | | | | Severe AKI | | | |
|---|---|---|---|---|---|---|---|---|---|---|---|---|---|
| | | 6 hrs | 12 hrs | 24 hrs | 48 hrs | 6 hrs | 12 hrs | 24 hrs | 48 hrs | 6 hrs | 12 hrs | 24 hrs | 48 hrs |
| **Gender** | Male | 0.904 | 0.901 | 0.9 | 0.89 | 0.944 | 0.95 | 0.942 | 0.923 | 0.982 | 0.979 | 0.977 | 0.96 |
| | Female | 0.885 | 0.882 | 0.874 | 0.877 | 0.919 | 0.923 | 0.918 | 0.899 | 0.955 | 0.949 | 0.956 | 0.931 |
| **Center** | The First Center | 0.899 | 0.896 | 0.891 | 0.883 | 0.944 | 0.935 | 0.935 | 0.917 | 0.972 | 0.969 | 0.968 | 0.943 |
| | The Third Center | 0.892 | 0.905 | 0.91 | 0.918 | 0.959 | 0.946 | 0.954 | 0.953 | 0.989 | 0.988 | 0.982 | 0.981 |
| **Age** | 18 - 35 | 0.907 | 0.89 | 0.9 | 0.899 | 0.943 | 0.943 | 0.947 | 0.941 | 0.987 | 0.982 | 0.98 | 0.975 |
| | 35 - 45 | 0.939 | 0.924 | 0.917 | 0.914 | 0.981 | 0.981 | 0.975 | 0.951 | 0.99 | 0.985 | 0.989 | 0.964 |
| | 45 - 55 | 0.901 | 0.906 | 0.908 | 0.907 | 0.953 | 0.943 | 0.949 | 0.949 | 0.978 | 0.974 | 0.976 | 0.974 |
| | 55 - 65 | 0.888 | 0.89 | 0.874 | 0.871 | 0.922 | 0.915 | 0.901 | 0.886 | 0.962 | 0.962 | 0.961 | 0.927 |
| | 65 - 75 | 0.898 | 0.9 | 0.885 | 0.868 | 0.945 | 0.94 | 0.929 | 0.906 | 0.953 | 0.955 | 0.955 | 0.93 |
| | 75 - 100 | 0.938 | 0.903 | 0.894 | 0.881 | 0.968 | 0.954 | 0.937 | 0.924 | 0.994 | 0.992 | 0.987 | 0.986 |
| **Year** | 2010 - 2012 | 0.904 | 0.901 | 0.893 | 0.883 | 0.95 | 0.95 | 0.943 | 0.928 | 0.983 | 0.984 | 0.983 | 0.95 |
| | 2012 - 2014 | 0.912 | 0.914 | 0.912 | 0.885 | 0.957 | 0.948 | 0.958 | 0.94 | 0.986 | 0.986 | 0.985 | 0.966 |
| | 2014 - 2016 | 0.902 | 0.894 | 0.897 | 0.883 | 0.93 | 0.935 | 0.928 | 0.904 | 0.981 | 0.98 | 0.978 | 0.957 |
| | 2016 - 2018 | 0.872 | 0.885 | 0.874 | 0.878 | 0.899 | 0.912 | 0.911 | 0.895 | 0.957 | 0.93 | 0.94 | 0.919 |
| | 2018 - 2020 | 0.909 | 0.899 | 0.889 | 0.882 | 0.947 | 0.936 | 0.938 | 0.909 | 0.959 | 0.964 | 0.956 | 0.949 |
| | 2020 - 2022 | 0.894 | 0.897 | 0.903 | 0.889 | 0.905 | 0.933 | 0.92 | 0.893 | 0.969 | 0.965 | 0.96 | 0.947 |
| **Surgery Type** | Artery Surgery | 0.903 | 0.894 | 0.914 | 0.895 | 0.925 | 0.941 | 0.935 | 0.928 | 0.953 | 0.961 | 0.953 | 0.928 |
| | Congenital Heart Surgery | 0.901 | 0.899 | 0.904 | 0.876 | 0.959 | 0.956 | 0.97 | 0.934 | 0.955 | 0.953 | 0.964 | 0.921 |
| | CABG | 0.903 | 0.895 | 0.891 | 0.88 | 0.953 | 0.947 | 0.945 | 0.928 | 0.99 | 0.989 | 0.984 | 0.968 |
| | Pericardiectomy | 0.913 | 0.912 | 0.896 | 0.892 | 0.904 | 0.887 | 0.855 | 0.842 | 0.989 | 0.993 | 0.992 | 0.953 |
| | Valve Surgery | 0.901 | 0.901 | 0.902 | 0.895 | 0.942 | 0.946 | 0.939 | 0.925 | 0.977 | 0.974 | 0.975 | 0.965 |
| | CABG+Valve Surgery | 0.864 | 0.897 | 0.861 | 0.865 | 0.912 | 0.907 | 0.899 | 0.902 | 0.975 | 0.916 | 0.961 | 0.909 |

| | | | | | | | | | | | | | |
|---|---|---|---|---|---|---|---|---|---|---|---|---|---|
| | Valve Surgery+Congenital Heart Surgery | 0.909 | 0.921 | 0.914 | 0.869 | 0.989 | 0.982 | 0.985 | 0.952 | 0.99 | 0.979 | 0.963 | 0.925 |
| | Valve Surgery+Artery Surgery | 0.767 | 0.866 | 0.844 | 0.824 | 0.431 | 0.436 | 0.602 | 0.692 | 0.472 | 0.631 | 0.651 | 0.749 |
| | Others Surgery | 0.872 | 0.878 | 0.85 | 0.856 | 0.936 | 0.914 | 0.894 | 0.883 | 0.966 | 0.918 | 0.934 | 0.905 |
| **Mode of admission** | Normal | 0.903 | 0.895 | 0.893 | 0.883 | 0.936 | 0.941 | 0.932 | 0.915 | 0.974 | 0.968 | 0.97 | 0.948 |
| | Emergency | 0.923 | 0.901 | 0.938 | 0.883 | 0.974 | 0.971 | 0.983 | 0.974 | 0.953 | 0.964 | 0.975 | 0.981 |

*CABG, coronary artery bypass grafting.*

### *I. The AUPRC of REACT at different time points in different subgroups in internal validation.*

| | | Mild AKI | | | | Moderate AKI | | | | Severe AKI | | | |
|---|---|---|---|---|---|---|---|---|---|---|---|---|---|
| | | 6 hrs | 12 hrs | 24 hrs | 48 hrs | 6 hrs | 12 hrs | 24 hrs | 48 hrs | 6 hrs | 12 hrs | 24 hrs | 48 hrs |
| **Gender** | Male | 0.695 | 0.682 | 0.672 | 0.626 | 0.725 | 0.733 | 0.695 | 0.645 | 0.808 | 0.801 | 0.799 | 0.735 |
| | Female | 0.589 | 0.588 | 0.579 | 0.557 | 0.598 | 0.59 | 0.595 | 0.522 | 0.608 | 0.602 | 0.587 | 0.557 |
| **Center** | The First Center | 0.666 | 0.662 | 0.64 | 0.595 | 0.695 | 0.668 | 0.665 | 0.605 | 0.731 | 0.719 | 0.725 | 0.655 |
| | The Third Center | 0.602 | 0.626 | 0.618 | 0.624 | 0.639 | 0.654 | 0.706 | 0.69 | 0.797 | 0.781 | 0.735 | 0.707 |
| **Age** | 18 - 35 | 0.702 | 0.691 | 0.684 | 0.62 | 0.837 | 0.81 | 0.779 | 0.702 | 0.814 | 0.868 | 0.773 | 0.759 |
| | 35 - 45 | 0.764 | 0.753 | 0.728 | 0.682 | 0.837 | 0.834 | 0.81 | 0.759 | 0.825 | 0.812 | 0.838 | 0.791 |
| | 45 - 55 | 0.601 | 0.63 | 0.618 | 0.593 | 0.694 | 0.703 | 0.698 | 0.617 | 0.668 | 0.743 | 0.759 | 0.714 |
| | 55 - 65 | 0.67 | 0.657 | 0.661 | 0.606 | 0.712 | 0.692 | 0.709 | 0.653 | 0.752 | 0.755 | 0.74 | 0.707 |
| | 65 - 75 | 0.697 | 0.689 | 0.653 | 0.621 | 0.687 | 0.646 | 0.649 | 0.597 | 0.665 | 0.68 | 0.683 | 0.659 |
| | 75 - 100 | 0.767 | 0.756 | 0.752 | 0.66 | 0.836 | 0.815 | 0.775 | 0.688 | 0.755 | 0.804 | 0.801 | 0.685 |
| **Year** | 2010 - 2012 | 0.647 | 0.647 | 0.625 | 0.573 | 0.651 | 0.637 | 0.622 | 0.562 | 0.696 | 0.722 | 0.709 | 0.584 |
| | 2012 - 2014 | 0.723 | 0.703 | 0.699 | 0.627 | 0.799 | 0.756 | 0.777 | 0.694 | 0.846 | 0.843 | 0.829 | 0.812 |
| | 2014 - 2016 | 0.692 | 0.677 | 0.693 | 0.648 | 0.741 | 0.714 | 0.719 | 0.658 | 0.751 | 0.726 | 0.739 | 0.691 |
| | 2016 - 2018 | 0.626 | 0.626 | 0.584 | 0.582 | 0.637 | 0.641 | 0.657 | 0.592 | 0.742 | 0.671 | 0.626 | 0.598 |
| | 2018 - 2020 | 0.686 | 0.689 | 0.651 | 0.609 | 0.721 | 0.689 | 0.672 | 0.589 | 0.694 | 0.725 | 0.681 | 0.696 |
| | 2020 - 2022 | 0.591 | 0.593 | 0.603 | 0.55 | 0.583 | 0.654 | 0.592 | 0.538 | 0.766 | 0.757 | 0.757 | 0.661 |
| **Surgery Type** | Artery Surgery | 0.671 | 0.676 | 0.708 | 0.631 | 0.656 | 0.711 | 0.685 | 0.626 | 0.676 | 0.718 | 0.674 | 0.623 |
| | Congenital Heart Surgery | 0.735 | 0.713 | 0.723 | 0.643 | 0.808 | 0.79 | 0.833 | 0.734 | 0.725 | 0.735 | 0.779 | 0.716 |
| | CABG | 0.689 | 0.673 | 0.656 | 0.6 | 0.751 | 0.716 | 0.701 | 0.647 | 0.82 | 0.831 | 0.805 | 0.727 |
| | Pericardiectomy | 0.713 | 0.685 | 0.636 | 0.623 | 0.588 | 0.602 | 0.435 | 0.45 | 0.552 | 0.772 | 0.534 | 0.484 |
| | Valve Surgery | 0.629 | 0.624 | 0.617 | 0.596 | 0.653 | 0.642 | 0.631 | 0.583 | 0.683 | 0.687 | 0.631 | 0.611 |
| | CABG+Valve Surgery | 0.57 | 0.641 | 0.542 | 0.539 | 0.689 | 0.644 | 0.593 | 0.529 | 0.843 | 0.715 | 0.754 | 0.617 |

| | | | | | | | | | | | | | |
|---|---|---|---|---|---|---|---|---|---|---|---|---|---|
| | Valve Surgery+Congenital Heart Surgery | 0.8 | 0.813 | 0.785 | 0.677 | 0.897 | 0.847 | 0.903 | 0.844 | 0.86 | 0.816 | 0.815 | 0.744 |
| | Valve Surgery+Artery Surgery | 0.264 | 0.461 | 0.306 | 0.24 | 0.009 | 0.008 | 0.015 | 0.03 | 0.014 | 0.014 | 0.008 | 0.041 |
| | Others Surgery | 0.674 | 0.684 | 0.609 | 0.583 | 0.778 | 0.701 | 0.692 | 0.64 | 0.924 | 0.823 | 0.863 | 0.726 |
| **Mode of admission** | Normal | 0.67 | 0.651 | 0.637 | 0.589 | 0.674 | 0.686 | 0.661 | 0.604 | 0.749 | 0.716 | 0.725 | 0.665 |
| | Emergency | 0.831 | 0.77 | 0.827 | 0.712 | 0.827 | 0.837 | 0.888 | 0.824 | 0.656 | 0.75 | 0.763 | 0.86 |

*CABG, coronary artery bypass grafting.*

### *J. The AUROC of REACT at different time points in different subgroups in external test.*

| | | Mild AKI | | | | Moderate AKI | | | | Severe AKI | | | |
|---|---|---|---|---|---|---|---|---|---|---|---|---|---|
| | | 6 hrs | 12 hrs | 24 hrs | 48 hrs | 6 hrs | 12 hrs | 24 hrs | 48 hrs | 6 hrs | 12 hrs | 24 hrs | 48 hrs |
| **Gender** | Male | 0.892 | 0.894 | 0.892 | 0.864 | 0.939 | 0.94 | 0.941 | 0.914 | 0.971 | 0.971 | 0.971 | 0.948 |
| | Female | 0.879 | 0.881 | 0.881 | 0.857 | 0.914 | 0.915 | 0.919 | 0.892 | 0.945 | 0.952 | 0.95 | 0.917 |
| **Center** | The Sixth Center | 0.839 | 0.838 | 0.825 | 0.808 | 0.906 | 0.9 | 0.899 | 0.875 | 0.951 | 0.944 | 0.945 | 0.906 |
| | The Seventh Center | 0.85 | 0.843 | 0.844 | 0.87 | 0.882 | 0.884 | 0.884 | 0.873 | 0.952 | 0.943 | 0.933 | 0.887 |
| | The Nangjing Drum | 0.891 | 0.891 | 0.893 | 0.861 | 0.933 | 0.934 | 0.936 | 0.906 | 0.966 | 0.966 | 0.966 | 0.94 |
| **Age** | 18 - 35 | 0.901 | 0.9 | 0.9 | 0.871 | 0.923 | 0.919 | 0.923 | 0.904 | 0.954 | 0.947 | 0.946 | 0.916 |
| | 35 - 45 | 0.917 | 0.916 | 0.919 | 0.894 | 0.942 | 0.945 | 0.947 | 0.928 | 0.97 | 0.973 | 0.971 | 0.952 |
| | 45 - 55 | 0.899 | 0.899 | 0.903 | 0.868 | 0.934 | 0.94 | 0.941 | 0.91 | 0.972 | 0.976 | 0.972 | 0.942 |
| | 55 - 65 | 0.878 | 0.88 | 0.882 | 0.851 | 0.933 | 0.934 | 0.939 | 0.907 | 0.966 | 0.967 | 0.965 | 0.938 |
| | 65 - 75 | 0.874 | 0.872 | 0.875 | 0.84 | 0.922 | 0.926 | 0.928 | 0.896 | 0.96 | 0.962 | 0.957 | 0.937 |
| | 75 - 100 | 0.858 | 0.859 | 0.856 | 0.825 | 0.893 | 0.901 | 0.894 | 0.865 | 0.955 | 0.944 | 0.944 | 0.913 |
| **Year** | 2010 - 2012 | 0.876 | 0.876 | 0.877 | 0.844 | 0.926 | 0.918 | 0.918 | 0.89 | 0.957 | 0.96 | 0.953 | 0.922 |
| | 2012 - 2014 | 0.862 | 0.862 | 0.863 | 0.844 | 0.908 | 0.903 | 0.911 | 0.886 | 0.956 | 0.958 | 0.954 | 0.92 |
| | 2014 - 2016 | 0.873 | 0.872 | 0.862 | 0.831 | 0.923 | 0.918 | 0.924 | 0.89 | 0.963 | 0.965 | 0.957 | 0.92 |
| | 2016 - 2018 | 0.881 | 0.885 | 0.887 | 0.859 | 0.931 | 0.932 | 0.927 | 0.901 | 0.957 | 0.958 | 0.957 | 0.928 |
| | 2018 - 2020 | 0.901 | 0.903 | 0.907 | 0.88 | 0.942 | 0.939 | 0.94 | 0.922 | 0.969 | 0.969 | 0.97 | 0.954 |
| | 2020 - 2022 | 0.884 | 0.884 | 0.886 | 0.852 | 0.931 | 0.933 | 0.934 | 0.902 | 0.967 | 0.965 | 0.967 | 0.937 |
| **Surgery Type** | Artery Surgery | 0.92 | 0.918 | 0.916 | 0.895 | 0.948 | 0.943 | 0.948 | 0.928 | 0.965 | 0.964 | 0.964 | 0.943 |
| | Congenital Heart Surgery | 0.844 | 0.853 | 0.863 | 0.82 | 0.907 | 0.895 | 0.903 | 0.879 | 0.967 | 0.963 | 0.961 | 0.954 |
| | CABG | 0.843 | 0.847 | 0.846 | 0.818 | 0.926 | 0.932 | 0.937 | 0.9 | 0.959 | 0.965 | 0.961 | 0.935 |
| | Pericardiectomy | 0.849 | 0.834 | 0.831 | 0.788 | 0.882 | 0.895 | 0.876 | 0.844 | 0.907 | 0.931 | 0.923 | 0.886 |
| | Valve Surgery | 0.877 | 0.873 | 0.87 | 0.849 | 0.915 | 0.911 | 0.913 | 0.877 | 0.942 | 0.946 | 0.937 | 0.902 |
| | CABG+Artery Surgery | 0.869 | 0.879 | 0.883 | 0.856 | 0.924 | 0.925 | 0.925 | 0.907 | 0.935 | 0.95 | 0.945 | 0.917 |

| | | | | | | | | | | | | | |
|---|---|---|---|---|---|---|---|---|---|---|---|---|---|
| | CABG+Valve Surgery | 0.882 | 0.881 | 0.881 | 0.855 | 0.917 | 0.917 | 0.922 | 0.897 | 0.967 | 0.963 | 0.965 | 0.936 |
| | Valve Surgery+Congenital Heart Surgery | 0.876 | 0.873 | 0.889 | 0.835 | 0.921 | 0.902 | 0.925 | 0.881 | 0.972 | 0.975 | 0.986 | 0.961 |
| | Valve Surgery+Artery Surgery | 0.892 | 0.889 | 0.895 | 0.86 | 0.947 | 0.942 | 0.941 | 0.919 | 0.982 | 0.979 | 0.982 | 0.966 |
| | Others Surgery | 0.895 | 0.89 | 0.899 | 0.877 | 0.936 | 0.944 | 0.94 | 0.91 | 0.973 | 0.976 | 0.979 | 0.966 |
| **Mode of admission** | Normal | 0.874 | 0.874 | 0.875 | 0.844 | 0.923 | 0.923 | 0.925 | 0.892 | 0.957 | 0.959 | 0.959 | 0.926 |
| | Emergency | 0.908 | 0.907 | 0.908 | 0.891 | 0.924 | 0.926 | 0.926 | 0.907 | 0.95 | 0.953 | 0.951 | 0.929 |

*CABG, coronary artery bypass grafting.*

#### *K. The AUPRC of REACT at different time points in different subgroups in external test.*

| | | Mild AKI | | | | Moderate AKI | | | | Severe AKI | | | |
|---|---|---|---|---|---|---|---|---|---|---|---|---|---|
| | | 6 hrs | 12 hrs | 24 hrs | 48 hrs | 6 hrs | 12 hrs | 24 hrs | 48 hrs | 6 hrs | 12 hrs | 24 hrs | 48 hrs |
| **Gender** | Male | 0.687 | 0.687 | 0.687 | 0.63 | 0.712 | 0.717 | 0.713 | 0.647 | 0.719 | 0.726 | 0.725 | 0.655 |
| | Female | 0.599 | 0.597 | 0.601 | 0.546 | 0.589 | 0.588 | 0.58 | 0.521 | 0.539 | 0.555 | 0.556 | 0.502 |
| **Center** | The Sixth Center | 0.606 | 0.607 | 0.592 | 0.554 | 0.596 | 0.595 | 0.601 | 0.55 | 0.593 | 0.582 | 0.581 | 0.523 |
| | The Seventh Center | 0.579 | 0.546 | 0.543 | 0.528 | 0.627 | 0.612 | 0.613 | 0.556 | 0.765 | 0.76 | 0.719 | 0.637 |
| | The Nangjing Drum | 0.661 | 0.662 | 0.667 | 0.599 | 0.68 | 0.674 | 0.677 | 0.611 | 0.675 | 0.683 | 0.674 | 0.611 |
| **Age** | 18 – 35 | 0.705 | 0.702 | 0.701 | 0.633 | 0.739 | 0.714 | 0.714 | 0.666 | 0.755 | 0.739 | 0.746 | 0.663 |
| | 35 – 45 | 0.758 | 0.752 | 0.756 | 0.707 | 0.74 | 0.75 | 0.749 | 0.709 | 0.736 | 0.742 | 0.735 | 0.676 |
| | 45 – 55 | 0.693 | 0.688 | 0.699 | 0.619 | 0.724 | 0.734 | 0.732 | 0.649 | 0.734 | 0.726 | 0.721 | 0.663 |
| | 55 – 65 | 0.616 | 0.621 | 0.625 | 0.558 | 0.662 | 0.66 | 0.654 | 0.587 | 0.641 | 0.645 | 0.65 | 0.581 |
| | 65 – 75 | 0.618 | 0.622 | 0.634 | 0.568 | 0.625 | 0.628 | 0.644 | 0.572 | 0.634 | 0.647 | 0.628 | 0.585 |
| | 75 – 100 | 0.622 | 0.621 | 0.619 | 0.569 | 0.542 | 0.561 | 0.54 | 0.491 | 0.512 | 0.505 | 0.486 | 0.429 |
| **Year** | 2010 – 2012 | 0.681 | 0.682 | 0.674 | 0.624 | 0.68 | 0.649 | 0.66 | 0.618 | 0.656 | 0.652 | 0.65 | 0.595 |
| | 2012 – 2014 | 0.61 | 0.61 | 0.613 | 0.569 | 0.607 | 0.598 | 0.61 | 0.545 | 0.575 | 0.575 | 0.588 | 0.524 |
| | 2014 – 2016 | 0.628 | 0.621 | 0.603 | 0.545 | 0.637 | 0.636 | 0.641 | 0.57 | 0.634 | 0.653 | 0.613 | 0.573 |
| | 2016 – 2018 | 0.661 | 0.659 | 0.659 | 0.593 | 0.673 | 0.678 | 0.665 | 0.597 | 0.699 | 0.703 | 0.694 | 0.614 |
| | 2018 – 2020 | 0.712 | 0.715 | 0.726 | 0.668 | 0.726 | 0.723 | 0.725 | 0.677 | 0.709 | 0.721 | 0.724 | 0.68 |
| | 2020 – 2022 | 0.631 | 0.625 | 0.629 | 0.56 | 0.65 | 0.661 | 0.657 | 0.586 | 0.666 | 0.648 | 0.662 | 0.579 |
| **Surgery Type** | Artery Surgery | 0.81 | 0.809 | 0.806 | 0.76 | 0.804 | 0.796 | 0.801 | 0.756 | 0.767 | 0.75 | 0.753 | 0.701 |
| | Congenital Heart Surgery | 0.513 | 0.538 | 0.558 | 0.459 | 0.52 | 0.533 | 0.531 | 0.479 | 0.506 | 0.484 | 0.467 | 0.517 |
| | CABG | 0.535 | 0.539 | 0.545 | 0.479 | 0.561 | 0.576 | 0.59 | 0.51 | 0.58 | 0.568 | 0.566 | 0.481 |
| | Pericardiectomy | 0.599 | 0.596 | 0.58 | 0.539 | 0.472 | 0.532 | 0.499 | 0.43 | 0.492 | 0.435 | 0.448 | 0.483 |
| | Valve Surgery | 0.578 | 0.568 | 0.566 | 0.522 | 0.591 | 0.581 | 0.59 | 0.51 | 0.601 | 0.612 | 0.574 | 0.507 |
| | CABG+Artery Surgery | 0.66 | 0.665 | 0.66 | 0.621 | 0.642 | 0.669 | 0.656 | 0.608 | 0.65 | 0.612 | 0.584 | 0.528 |

| | | | | | | | | | | | | | |
|---|---|---|---|---|---|---|---|---|---|---|---|---|---|
| | CABG+Valve Surgery | 0.613 | 0.614 | 0.625 | 0.583 | 0.547 | 0.572 | 0.578 | 0.539 | 0.656 | 0.672 | 0.656 | 0.617 |
| | Valve Surgery+Congenital Heart Surgery | 0.498 | 0.542 | 0.562 | 0.439 | 0.502 | 0.466 | 0.509 | 0.438 | 0.632 | 0.615 | 0.64 | 0.564 |
| | Valve Surgery+Artery Surgery | 0.556 | 0.565 | 0.584 | 0.517 | 0.602 | 0.61 | 0.604 | 0.553 | 0.589 | 0.596 | 0.622 | 0.572 |
| | Others Surgery | 0.598 | 0.625 | 0.648 | 0.599 | 0.624 | 0.664 | 0.672 | 0.581 | 0.758 | 0.752 | 0.766 | 0.72 |
| **Mode of admission** | Normal | 0.588 | 0.586 | 0.591 | 0.527 | 0.605 | 0.602 | 0.607 | 0.543 | 0.627 | 0.632 | 0.627 | 0.56 |
| | Emergency | 0.816 | 0.815 | 0.816 | 0.784 | 0.773 | 0.778 | 0.777 | 0.738 | 0.747 | 0.747 | 0.732 | 0.688 |

*CABG, coronary artery bypass grafting.*

#### *L. The number of input variables and the model performance of REACT with different λ.*

| **Λ** | **Variable num** | **Mild AKI** 6 hrs | 12 hrs | 24 hrs | 48 hrs | **Moderate AKI** 6 hrs | 12 hrs | 24 hrs | 48 hrs | **Severe AKI** 6 hrs | 12 hrs | 24 hrs | 48 hrs |
|---|---|---|---|---|---|---|---|---|---|---|---|---|---|
| **AUROC** | | | | | | | | | | | | | |
| 5e-05 | 1 | 0.88 /0.82 | 0.88 /0.82 | 0.88 /0.82 | 0.88 /0.78 | 0.91 /0.86 | 0.91 /0.86 | 0.90 /0.86 | 0.89 /0.82 | 0.93 /0.91 | 0.93 /0.91 | 0.92 /0.91 | 0.91 /0.88 |
| 5e-04 | 6 | 0.89 /0.84 | 0.89 /0.84 | 0.89 /0.84 | 0.89 /0.80 | 0.92 /0.88 | 0.92 /0.89 | 0.92 /0.89 | 0.91 /0.85 | 0.95 /0.93 | 0.95 /0.93 | 0.95 /0.93 | 0.94 /0.90 |
| 5e-03 | 31 | 0.90 /0.89 | 0.90 /0.89 | 0.89 /0.89 | 0.88 /0.86 | 0.94 /0.93 | 0.94 /0.93 | 0.94 /0.93 | 0.92 /0.90 | 0.97 /0.96 | 0.97 /0.96 | 0.97 /0.96 | 0.95 /0.94 |
| 5e-02 | 59 | 0.84 /0.80 | 0.84 /0.80 | 0.83 /0.81 | 0.83 /0.78 | 0.88 /0.84 | 0.88 /0.84 | 0.88 /0.84 | 0.86 /0.81 | 0.91 /0.87 | 0.91 /0.87 | 0.91 /0.87 | 0.89 /0.84 |
| **AUPRC** | | | | | | | | | | | | | |
| 5e-05 | 1 | 0.82 /0.61 | 0.82 /0.61 | 0.82 /0.59 | 0.78 /0.57 | 0.86 /0.57 | 0.86 /0.57 | 0.86 /0.55 | 0.82 /0.50 | 0.91 /0.61 | 0.91 /0.60 | 0.91 /0.58 | 0.88 /0.53 |
| 5e-04 | 6 | 0.84 /0.62 | 0.84 /0.62 | 0.84 /0.60 | 0.80 /0.57 | 0.88 /0.60 | 0.89 /0.59 | 0.89 /0.58 | 0.85 /0.52 | 0.93 /0.64 | 0.93 /0.63 | 0.93 /0.61 | 0.90 /0.56 |
| 5e-03 | 31 | 0.89 /0.66 | 0.89 /0.65 | 0.89 /0.64 | 0.86 /0.60 | 0.93 /0.69 | 0.93 /0.68 | 0.93 /0.67 | 0.90 /0.61 | 0.96 /0.74 | 0.96 /0.74 | 0.96 /0.72 | 0.94 /0.66 |
| 5e-02 | 59 | 0.80 /0.48 | 0.80 /0.47 | 0.81 /0.46 | 0.78 /0.43 | 0.84 /0.44 | 0.84 /0.43 | 0.84 /0.41 | 0.81 /0.37 | 0.87 /0.39 | 0.87 /0.38 | 0.87 /0.36 | 0.84 /0.33 |

*The notation "/" separates the performance metrics of our model on external test sets (before) from its performance on internal validation sets (behind).*

### *M. The sensitivity (True Positive Rate) corresponding to different CSA-AKI risk score thresholds.*

| **Threshold** | **Mild AKI** | | | | **Moderate AKI** | | | | **Severe AKI** | | | |
|---|---|---|---|---|---|---|---|---|---|---|---|---|
| | 6 hrs | 12 hrs | 24 hrs | 48 hrs | 6 hrs | 12 hrs | 24 hrs | 48 hrs | 6 hrs | 12 hrs | 24 hrs | 48hrs |
| 0.01 | 0.97 | 0.98 | 0.98 | 0.97 | 0.94 | 0.94 | 0.95 | 0.95 | 0.95 | 0.95 | 0.95 | 0.93 |
| 0.02 | 0.95 | 0.96 | 0.96 | 0.95 | 0.91 | 0.91 | 0.92 | 0.91 | 0.93 | 0.92 | 0.93 | 0.87 |
| 0.05 | 0.91 | 0.92 | 0.91 | 0.90 | 0.85 | 0.85 | 0.86 | 0.81 | 0.86 | 0.86 | 0.88 | 0.82 |
| 0.1 | 0.85 | 0.86 | 0.85 | 0.82 | 0.77 | 0.78 | 0.79 | 0.72 | 0.80 | 0.80 | 0.82 | 0.76 |
| 0.2 | 0.75 | 0.77 | 0.75 | 0.67 | 0.68 | 0.68 | 0.69 | 0.61 | 0.71 | 0.72 | 0.74 | 0.68 |
| 0.3 | 0.67 | 0.68 | 0.67 | 0.57 | 0.61 | 0.61 | 0.62 | 0.54 | 0.64 | 0.65 | 0.68 | 0.61 |
| 0.4 | 0.60 | 0.61 | 0.60 | 0.49 | 0.56 | 0.56 | 0.58 | 0.49 | 0.58 | 0.59 | 0.62 | 0.54 |
| 0.5 | 0.54 | 0.54 | 0.53 | 0.42 | 0.53 | 0.52 | 0.53 | 0.44 | 0.53 | 0.55 | 0.56 | 0.48 |
| 0.6 | 0.47 | 0.47 | 0.47 | 0.35 | 0.49 | 0.47 | 0.49 | 0.38 | 0.48 | 0.49 | 0.51 | 0.42 |
| 0.7 | 0.40 | 0.39 | 0.40 | 0.29 | 0.43 | 0.42 | 0.43 | 0.32 | 0.43 | 0.44 | 0.44 | 0.35 |
| 0.8 | 0.33 | 0.32 | 0.33 | 0.22 | 0.37 | 0.35 | 0.35 | 0.25 | 0.37 | 0.39 | 0.39 | 0.28 |
| 0.9 | 0.23 | 0.23 | 0.24 | 0.13 | 0.27 | 0.26 | 0.26 | 0.16 | 0.26 | 0.28 | 0.27 | 0.17 |

### *N. The specificity (True Negative Rate) corresponding to different CSA-AKI risk score thresholds.*

| **Threshold** | **Mild AKI** | | | | **Moderate AKI** | | | | **Severe AKI** | | | |
|---|---|---|---|---|---|---|---|---|---|---|---|---|
| | 6 hrs | 12 hrs | 24 hrs | 48 hrs | 6 hrs | 12 hrs | 24 hrs | 48 hrs | 6 hrs | 12 hrs | 24 hrs | 48hrs |
| 0.01 | 0.29 | 0.26 | 0.27 | 0.25 | 0.66 | 0.65 | 0.65 | 0.50 | 0.84 | 0.86 | 0.83 | 0.73 |
| 0.02 | 0.42 | 0.40 | 0.39 | 0.38 | 0.78 | 0.77 | 0.77 | 0.65 | 0.90 | 0.91 | 0.90 | 0.86 |
| 0.05 | 0.59 | 0.57 | 0.59 | 0.54 | 0.88 | 0.88 | 0.88 | 0.85 | 0.95 | 0.95 | 0.95 | 0.94 |
| 0.1 | 0.75 | 0.73 | 0.75 | 0.70 | 0.93 | 0.93 | 0.93 | 0.93 | 0.97 | 0.97 | 0.97 | 0.96 |
| 0.2 | 0.87 | 0.86 | 0.87 | 0.87 | 0.97 | 0.97 | 0.96 | 0.97 | 0.98 | 0.98 | 0.98 | 0.98 |
| 0.3 | 0.92 | 0.91 | 0.92 | 0.93 | 0.98 | 0.98 | 0.98 | 0.98 | 0.99 | 0.99 | 0.98 | 0.99 |
| 0.4 | 0.94 | 0.94 | 0.95 | 0.96 | 0.98 | 0.98 | 0.98 | 0.99 | 0.99 | 0.99 | 0.99 | 0.99 |
| 0.5 | 0.96 | 0.96 | 0.97 | 0.98 | 0.99 | 0.99 | 0.99 | 0.99 | 0.99 | 0.99 | 0.99 | 0.99 |
| 0.6 | 0.98 | 0.98 | 0.98 | 0.99 | 0.99 | 0.99 | 0.99 | 0.99 | 1.00 | 0.99 | 0.99 | 1.00 |
| 0.7 | 0.99 | 0.99 | 0.99 | 0.99 | 0.99 | 0.99 | 0.99 | 1.00 | 1.00 | 1.00 | 1.00 | 1.00 |
| 0.8 | 0.99 | 0.99 | 0.99 | 1.00 | 0.99 | 1.00 | 1.00 | 1.00 | 1.00 | 1.00 | 1.00 | 1.00 |
| 0.9 | 1.00 | 1.00 | 1.00 | 1.00 | 1.00 | 1.00 | 1.00 | 1.00 | 1.00 | 1.00 | 1.00 | 0.73 |

***O. The False discovery rate corresponding to different CSA-AKI risk score thresholds.***

| **Threshold** | **Mild AKI** | | | | **Moderate AKI** | | | | **Severe AKI** | | | |
|---|---|---|---|---|---|---|---|---|---|---|---|---|
| | 6 hrs | 12 hrs | 24 hrs | 48 hrs | 6 hrs | 12 hrs | 24 hrs | 48 hrs | 6 hrs | 12 hrs | 24 hrs | 48hrs |
| 0.01 | 0.85 | 0.85 | 0.85 | 0.85 | 0.85 | 0.85 | 0.85 | 0.89 | 0.85 | 0.83 | 0.85 | 0.91 |
| 0.02 | 0.82 | 0.83 | 0.83 | 0.83 | 0.79 | 0.79 | 0.79 | 0.86 | 0.77 | 0.75 | 0.78 | 0.84 |
| 0.05 | 0.77 | 0.78 | 0.77 | 0.79 | 0.68 | 0.69 | 0.69 | 0.75 | 0.64 | 0.63 | 0.66 | 0.72 |
| 0.1 | 0.69 | 0.70 | 0.69 | 0.73 | 0.57 | 0.58 | 0.58 | 0.62 | 0.55 | 0.54 | 0.57 | 0.62 |
| 0.2 | 0.57 | 0.58 | 0.56 | 0.59 | 0.42 | 0.43 | 0.44 | 0.46 | 0.46 | 0.46 | 0.48 | 0.50 |
| 0.3 | 0.48 | 0.49 | 0.48 | 0.47 | 0.35 | 0.35 | 0.36 | 0.37 | 0.40 | 0.41 | 0.43 | 0.44 |
| 0.4 | 0.41 | 0.41 | 0.40 | 0.38 | 0.31 | 0.30 | 0.32 | 0.32 | 0.35 | 0.36 | 0.37 | 0.38 |
| 0.5 | 0.34 | 0.34 | 0.33 | 0.31 | 0.28 | 0.27 | 0.28 | 0.27 | 0.30 | 0.31 | 0.32 | 0.32 |
| 0.6 | 0.27 | 0.27 | 0.26 | 0.23 | 0.25 | 0.24 | 0.25 | 0.23 | 0.25 | 0.26 | 0.28 | 0.27 |
| 0.7 | 0.20 | 0.19 | 0.20 | 0.17 | 0.21 | 0.20 | 0.21 | 0.19 | 0.21 | 0.23 | 0.23 | 0.20 |
| 0.8 | 0.14 | 0.14 | 0.14 | 0.12 | 0.17 | 0.16 | 0.16 | 0.13 | 0.17 | 0.18 | 0.18 | 0.14 |
| 0.9 | 0.10 | 0.10 | 0.10 | 0.06 | 0.11 | 0.11 | 0.11 | 0.07 | 0.13 | 0.13 | 0.13 | 0.09 |

### *P. The recommend thresholds and its corresponding evaluation indexes.*

| | | Mild AKI | | | | Moderate AKI | | | | Severe AKI | | | |
|---|---|---|---|---|---|---|---|---|---|---|---|---|---|
| | | 6 hrs | 12 hrs | 24 hrs | 48 hrs | 6 hrs | 12 hrs | 24 hrs | 48 hrs | 6 hrs | 12 hrs | 24 hrs | 48hrs |
| **External tests** | Threshold | 0.11 | 0.12 | 0.11 | 0.14 | 0.05 | 0.06 | 0.06 | 0.07 | 0.04 | 0.04 | 0.05 | 0.07 |
| | TPR | 0.83 | 0.83 | 0.83 | 0.75 | 0.84 | 0.84 | 0.85 | 0.76 | 0.88 | 0.88 | 0.88 | 0.79 |
| | FDR | 0.67 | 0.67 | 0.67 | 0.67 | 0.67 | 0.67 | 0.67 | 0.67 | 0.67 | 0.67 | 0.67 | 0.67 |
| | TNR | 0.77 | 0.77 | 0.78 | 0.80 | 0.89 | 0.89 | 0.89 | 0.90 | 0.94 | 0.94 | 0.95 | 0.95 |
| **Internal Validation** | Threshold | 0.09 | 0.10 | 0.10 | 0.12 | 0.04 | 0.04 | 0.04 | 0.07 | 0.02 | 0.02 | 0.03 | 0.05 |
| | TPR | 0.84 | 0.83 | 0.82 | 0.80 | 0.82 | 0.82 | 0.81 | 0.71 | 0.88 | 0.88 | 0.86 | 0.78 |
| | FDR | 0.67 | 0.67 | 0.67 | 0.67 | 0.67 | 0.67 | 0.67 | 0.67 | 0.67 | 0.67 | 0.67 | 0.67 |
| | TNR | 0.78 | 0.79 | 0.80 | 0.80 | 0.91 | 0.92 | 0.92 | 0.93 | 0.95 | 0.95 | 0.96 | 0.96 |

*TPR, True Positive Rate, sensitivity; FDR, False Discovery Rate; TNR, True Negative Rate, specificity*.

***Q. Comparison of patient characteristics on cohorts whether AKI stage I is observed by doctors.***

| | **Observed** (n = 4,939) | **Unobserved** (n = 842) | **Unoccurred** (n = 23,234) | ***P-value*** |
|---|---|---|---|---|
| Sex (%) | | | | 0.003 |
| Female | 1883 (38.1) | 304 (36.1) | 9385 (40.4) | |
| Male | 3056 (61.9) | 538 (63.9) | 13840 (59.6) | |
| Age (median [IQR]) | 63.00 [53.00, 70.00] | 56.00 [46.00, 66.00] | 57.00 [47.00, 66.00] | <0.001 |
| **Center** | | | | <0.001 |
| The First Center | 2177 (44.1) | 220 (26.1) | 8978 (38.6) | |
| The Third Center | 162 (3.3) | 74 (8.8) | 709 (3.1) | |
| The Sixth Center | 368 (7.5) | 44 (5.2) | 1524 (6.6) | |
| The Seventh Center | 71 (1.4) | 16 (1.9) | 239 (1.0) | |
| The Nangjing Drum | 2161 (43.8) | 488 (58.0) | 11784 (50.7) | |
| **Commodities** | | | | |
| Hypertension | 3639 (73.7) | 632 (75.1) | 12874 (55.4) | <0.001 |
| Diabetes | 1536 (31.1) | 191 (22.7) | 4688 (20.2) | <0.001 |
| Congestive heart failure | 2144 (43.4) | 359 (42.6) | 9731 (41.9) | 0.137 |
| Pulmonary disease | 1215 (24.6) | 219 (26.0) | 4928 (21.2) | <0.001 |
| Chronic kidney disease | 484 (9.8) | 216 (25.7) | 522 (2.2) | <0.001 |
| **Outcome** | | | | |
| Death (%) | 236 (4.8) | 96 (11.4) | 97 (0.4) | <0.001 |
| Length of Stay (median [IQR]) | 22.86 [17.14, 32.02] | 24.00 [16.00, 35.00] | 19.04 [14.95, 26.00] | <0.001 |

### *R. Convergence conditions for Granger causality*

We show in Theorem 1 that under certain assumptions, the discovered causal vector will converge to the true Granger causal relationships.

**Assumption 1.** $\exists \lambda_0, \forall i = 1, \dots, N,\ \mathcal{L}_{\text{pred}}(\boldsymbol{Y}, \boldsymbol{X} \odot \boldsymbol{S}_{i=0}) - \mathcal{L}_{\text{pred}}(\boldsymbol{Y}, \boldsymbol{X} \odot \boldsymbol{S}_{i=1}) > \lambda_0$ if and only if time-series $i$ Granger cause $Y$, where $S_{i=l}$ is set S with element $s_i = l$.

**Theorem 1.** Given a time-series dataset $\langle \boldsymbol{X}, \boldsymbol{Y} \rangle$ and prediction model $f_\phi(\cdot)$. Under Assumptions 1, $\exists \lambda$, CPG element $m_i = \sigma(\theta_i)$ converges to 0 if time-series $i$ does not Granger cause $Y$, and $m_i$ converges to 1 if time-series i Granger cause $Y$.

The implications behind Assumption 1 can be intuitively explained. It means there exists a threshold $\lambda_0$ to binarize $\mathcal{L}_{\text{pred}}(\boldsymbol{Y}, \boldsymbol{X} \odot \boldsymbol{S}_{i=0}) - \mathcal{L}_{\text{pred}}(\boldsymbol{Y}, \boldsymbol{X} \odot \boldsymbol{S}_{i=1})$, serving as an indicator as to whether time-series $i$ contributes to prediction of $Y$. The proof of Theorem1 is provided in the supplementary material.

**Proof of Theorem 1:** In *Causal Discovery Stage* the loss function

$$\mathcal{L}_{\text{graph}}(\boldsymbol{X}, \boldsymbol{Y}, \boldsymbol{\theta}) = \mathcal{L}_{\text{pred}}(\boldsymbol{Y}, \boldsymbol{X} \odot \boldsymbol{S}) + \lambda \|\sigma(\boldsymbol{\theta})\|_1$$

where $s_i \sim Ber(\sigma(\theta_i))$. We use the REINFORCE [1] trick for theoretical analysis and $\theta_i$'s gradient is calculated as

$$\begin{aligned}\frac{\partial}{\partial \theta_{\mathrm{i}}} \mathbb{E}_{s_{\mathrm{i}}}[\mathcal{L}_{\text{graph}}] &= \mathbb{E}_{s_i}[\mathcal{L}_{\text{pred}}(\boldsymbol{Y}, \boldsymbol{X} \odot \boldsymbol{S})] + \lambda \sigma'(\theta_{\tau,ij}) \\ &= \mathcal{L}_{\text{pred}}(\boldsymbol{Y}, \boldsymbol{X} \odot \boldsymbol{S}_{i=1}) \sigma(\theta_i) \frac{\partial}{\partial \theta_{\mathrm{i}}} \sigma(\theta_i) + \mathcal{L}_{\text{pred}}(\boldsymbol{Y}, \boldsymbol{X} \odot \boldsymbol{S}_{i=0}) (1 - \sigma(\theta_i)) \frac{\partial}{\partial \theta_{\mathrm{i}}} (1 - \sigma(\theta_i)) + \lambda \sigma'(\theta_{\tau,ij}) \\ &= \sigma'(\theta_i) \big( \mathcal{L}_{\text{pred}}(\boldsymbol{Y}, \boldsymbol{X} \odot \boldsymbol{S}_{i=1}) - \mathcal{L}_{\text{pred}}(\boldsymbol{Y}, \boldsymbol{X} \odot \boldsymbol{S}_{i=0}) + \lambda \big)\end{aligned}$$

where $S_{i=l}$ is set S with element $s_i = l$, and $f_\phi(X \odot S) \equiv f_\phi(x_{t-\tau:t-1,1} \cdot s_1, \dots, x_{t-\tau:t-1,n} \cdot s_n)$, where $\tau$ is the maximal time lag. According to Definition 1, time-series $i$ does not Granger cause $Y$ if $x_{t-\tau:t-1,i}$ is invariant of the prediction of $Y$. Then we have $\forall c \in \{1, \dots, \mathrm{M}\}, f_{\phi,c}(\dots, x_{t-\tau,i}, \dots) = f_{\phi_j}(\dots, 0, \dots)$,

i.e., $\mathcal{L}_{\text{pred}}(\boldsymbol{Y}, \boldsymbol{X} \odot \boldsymbol{S}_{i=1}) = \mathcal{L}_{\text{pred}}(\boldsymbol{Y}, \boldsymbol{X} \odot \boldsymbol{S}_{i=0})$. Then we can derive that

$$\frac{\partial}{\partial \theta_{\mathrm{i}}} \mathbb{E}_{s_{\mathrm{i}}}[\mathcal{L}_{\text{graph}}] = \lambda \sigma'(\theta_i)$$

This is a sigmoidal gradient, whose convergence is analyzed in [2]. Likewise, we have $\exists c \in \{1, \dots, M\},\ f_{\phi,c}(\dots, x_{t-\tau,i}, \dots) \neq f_{\phi_j}(\dots, 0, \dots)$ if time-series $i$ does not Granger cause $Y$. With Assumption 1, $\exists \lambda_0, \forall i = 1, \dots, N,\ \mathcal{L}_{\text{pred}}(\boldsymbol{Y}, \boldsymbol{X} \odot \boldsymbol{S}_{i=0}) - \mathcal{L}_{\text{pred}}(\boldsymbol{Y}, \boldsymbol{X} \odot \boldsymbol{S}_{i=1}) > \lambda_0$. when we select the hyper-parameter $\lambda < \lambda_0$, we have

$$\frac{\partial}{\partial \theta_{\mathrm{i}}} \mathbb{E}_{s_{\mathrm{i}}}[\mathcal{L}_{\text{graph}}] = \sigma'(\theta_i) \big( \mathcal{L}_{\text{pred}}(\boldsymbol{Y}, \boldsymbol{X} \odot \boldsymbol{S}_{i=1}) - \mathcal{L}_{\text{pred}}(\boldsymbol{Y}, \boldsymbol{X} \odot \boldsymbol{S}_{i=0}) + \lambda \big) < \sigma'(\theta_i)(\lambda - \lambda_0) < 0$$

This gradient is expected to be negative, and $\theta_i$ will go towards $+\infty$ and $m_i \to 1$. In our implementation, we use Gumbel-Softmax estimator for improved performance and lower variance [3].

#### *S. Related works for causal discovery*

Our approach supports CSA-AKI prediction generalizable to different environments by identifying causal relationships, which is related to the area of causal discovery. Causal Discovery (or Causal Structural Learning), including static settings and dynamic time-series, has been a hot topic in machine learning and made big progress in the past decades. We roughly categorized these methods into multiple classes.

(i) **Constraint-based approaches:** such as PC [4], FCI[5], build causal graphs by performing conditional independence tests. This line of works are later extended to time-series settings, e.g. tsFCI [6], and PCMCI [7]–[9].

(ii) **Score-based learning algorithms:** include recovering causal graphs by applying penalized Neural Ordinary Differential Equations or acyclicity constraint [10], [11].

(iii) **Additive Noise Model (ANM):** infer causal graph based on additive noise assumption [12], under the assumptions of linear data generating process, no unobserved confounders, and disturbance variables have non-Gaussian distributions of non-zero variances. ANM is extended by [13] to nonlinear models with almost any nonlinearities and additive noise.

(iv) **Granger-causality-based approaches:** Granger causality was initially introduced by [14] who proposed to analyze the temporal causal relationships by testing the help of a time-series on predicting another time-series. Recently, Deep Neural Networks (NNs) have been widely applied to nonlinear Granger causality discovery [2], [15]–[19].

(v) **Convergent Cross Mapping (CCM):** proposed by [20] that reconstructs nonlinear state space for nonseparable weakly connected dynamic systems. This approach is later extended to situations of synchrony, confounding, or sporadic time-series [21]–[23].

#### *T. Implementation details*

We conduct experiments on a PC with Intel Core CPUs and NVIDIA GeForce RTX 3090 GPUs. We implement our algorithm and neural-network-based comparison algorithms with PyTorch[1], XGBoost with python package xgboost[2].

##### a) Network Architecture

Our causal learning model in REACT consists of two stages, outcomes prediction and causal discovery. The former stage utilize a neural network based on LSTM and MLP. To be able to handle both static and dynamic input, the network can be further divided into three parts: static encoder with MLP, dynamic encoder with LSTM, and decoder with another MLP.

As for comparison models, "LSTM / Transformer" in Fig. 3 means the same architecture with LSTM / Transformer dynamic encoder. "MLP" means all-MLP network, i.e., flatten all dynamic and static input before feeding into a single MLP encoder. "MLP", "LSTM", and "XGBoost" are without causal discovery, i.e., all variables are used in the prediction. In the experiments shown in Extended Figure 5, we show that training a regular neural network (e.g. MLP, LSTM, Transformer) with only selected variables, i.e., performing feature selection before deep learning instead of our causal learning strategy. For this setting, we only use the selected variables as input and set other variables to 0, which permit using the same network architecture with the same hidden layer width.

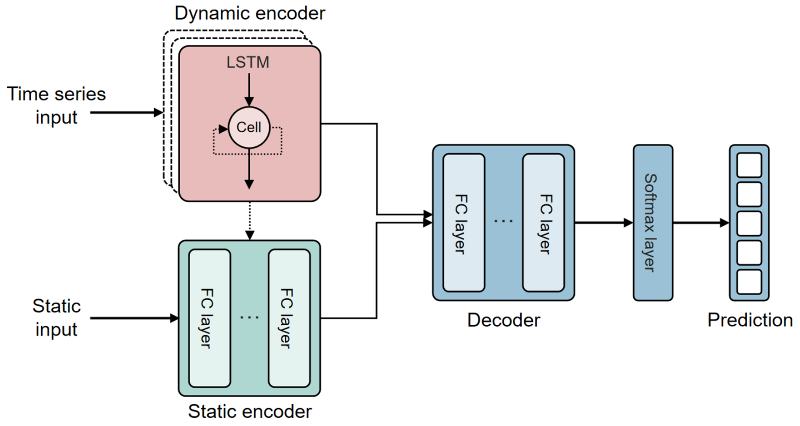


**Supplementary Figure 1.** Network architecture.

[1] https://pytorch.org/
[2] https://pypi.org/project/xgboost/

### b) Training Strategy

**Outcomes prediction stage.** The learning rate is set to be relatively high initially and gradually decrease to 10% of the initial value (using torch.optim.lr_scheduler.StepLR, $\gamma = 0.1^{1/\mathrm{tot\_epoch}}$).

**Causal discovery stage.** Initially, each element in the causal probability graph $\sigma(\boldsymbol{\theta})$ is set to be 0.5, i.e. $\boldsymbol{\theta}$ being all 0. The learning rate for this stage is set differently from the outcomes prediction stage. The learning rate also gradually decrease to 10% of the initial value. See the detailed settings of the parameters in the following section. To avoid the $\|\sigma(\boldsymbol{\theta})\|_1$ from being too large, it is first divided by the total numbers of the elements in $\boldsymbol{\theta}$ and then multiplied by $\lambda$. As a result

**XGBoost.** Our dataset contains more than a million samples, which is too large to load into memory. We implement XGBoost with data iterator, each containing 50000 samples.

### c) Parameter Search

Hyperparameters play important roles in deep learning model developments. We perform standard parameter grid search for each of the deep learning models and our causal learning model. The best set of hyperparameters is selected by choosing the best performance on the validation dataset. Since each training takes hours or even days, we can only covers a few values in the grid search, which is demonstrated as follow.

| **Parameters** | **Ours** | **MLP** | **LSTM** | |
|---|---|---|---|---|
| Batch size | 512, 1024, 2048 | 512, 1024, 2048 | 512, 1024, 2048 | / |
| Total epoch number | 10, 20, 40 | 10, 20, 40 | 10, 20, 40 | / |
| Activation | LeakyReLU | LeakyReLU | LeakyReLU | / |
| Encoder layer | 3, 6 | 3, 6 | 3, 6 | / |
| Decoder layer | 3, 6 | 3, 6 | 3, 6 | / |
| Hidden width | 32, 128 | 32, 128 | 32, 128 | / |

| | | | | |
|---|---|---|---|---|
| Weight decay | $0,\ 10^{-4},\ 3\times10^{-4},\ 10^{-3}$ | $0,\ 10^{-4},\ 3\times10^{-4},\ 10^{-3}$ | $0,\ 10^{-4},\ 3\times10^{-4},\ 10^{-3}$ | / |
| Dropout | 0, 0.1, 0.25 | 0, 0.1, 0.25 | 0, 0.1, 0.25 | / |
| τ | 0.1, 0.01 | 0.1, 0.01 | 0.1, 0.01 | / |
| λ | $5\times10^{-5},\ 5\times10^{-4},\ 5\times10^{-3},\ 5\times10^{-2}$ | $5\times10^{-5},\ 5\times10^{-4},\ 5\times10^{-3},\ 5\times10^{-2}$ | $5\times10^{-5},\ 5\times10^{-4},\ 5\times10^{-3},\ 5\times10^{-2}$ | / |

### U. *More Examples with Counterfactual Explanations*

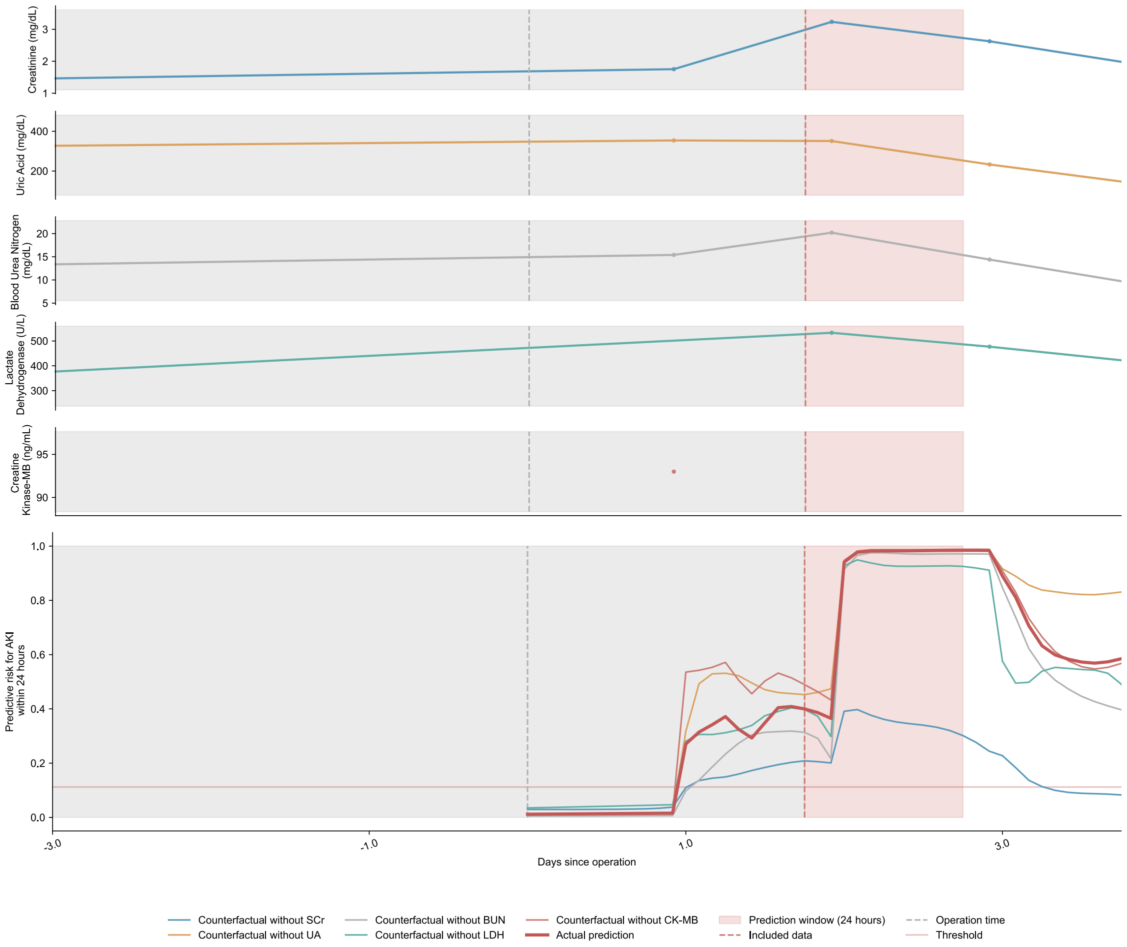


**Supplementary Figure U-1.** A female patient in their 50s with hypertension, diabetes mellitus, and chronic heart failure was admitted to the hospital for coronary (aortic) coronary artery bypass grafting, mitral-valve replacement, and tricuspid valvuloplasty. The length of stay of the patients was 37 days. Creatinine, uric acid, and urea nitrogen indices were stable before surgery, with a slight increase in creatine kinase enzymes. Finally, the patient developed CSA-AKI.

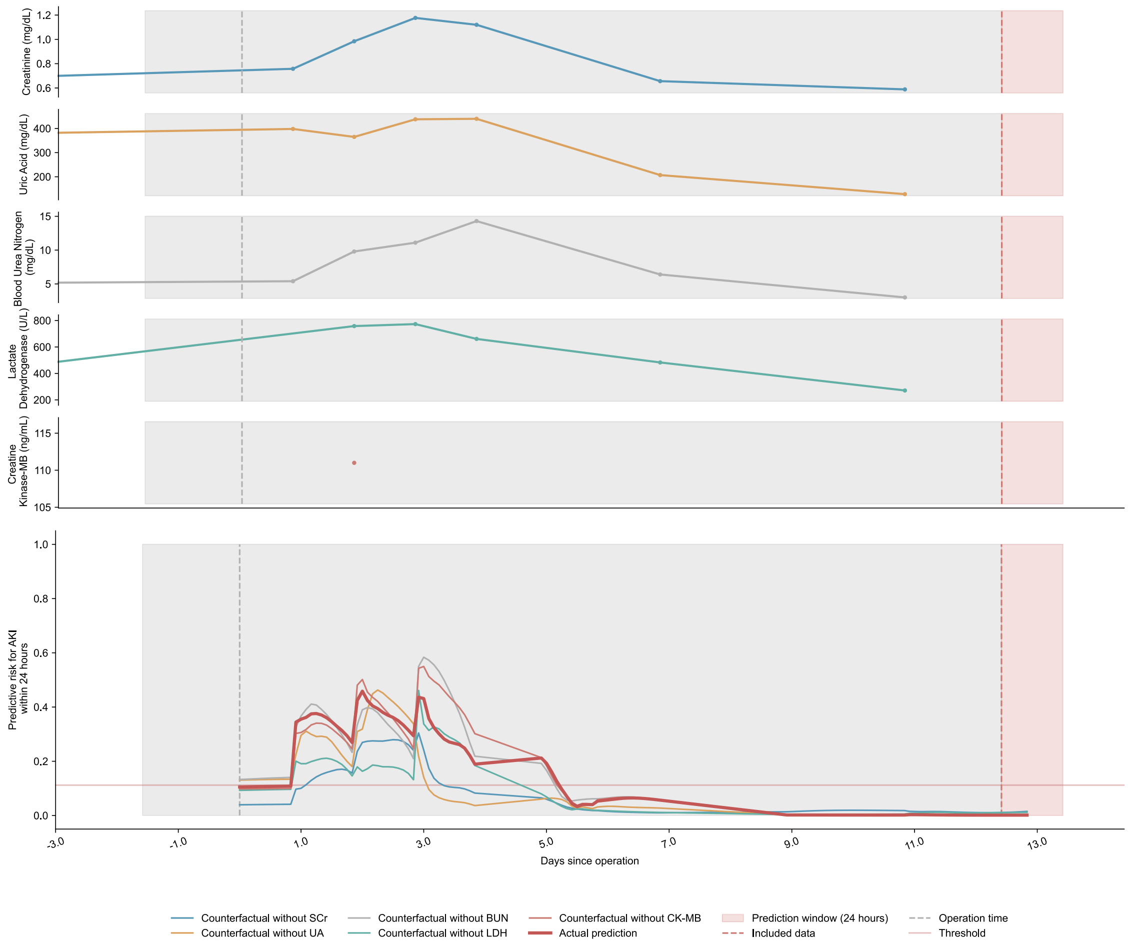


**Supplementary Figure U-2.** A lengthy 38-day admission of a male in their 50s with a history of hypertension and diabetes. The patient underwent atrial septal defect repair combined with tricuspid valvuloplasty. Although creatinine, uric acid and urea nitrogen were slightly increased after surgery, no CSA-AKI occurred.

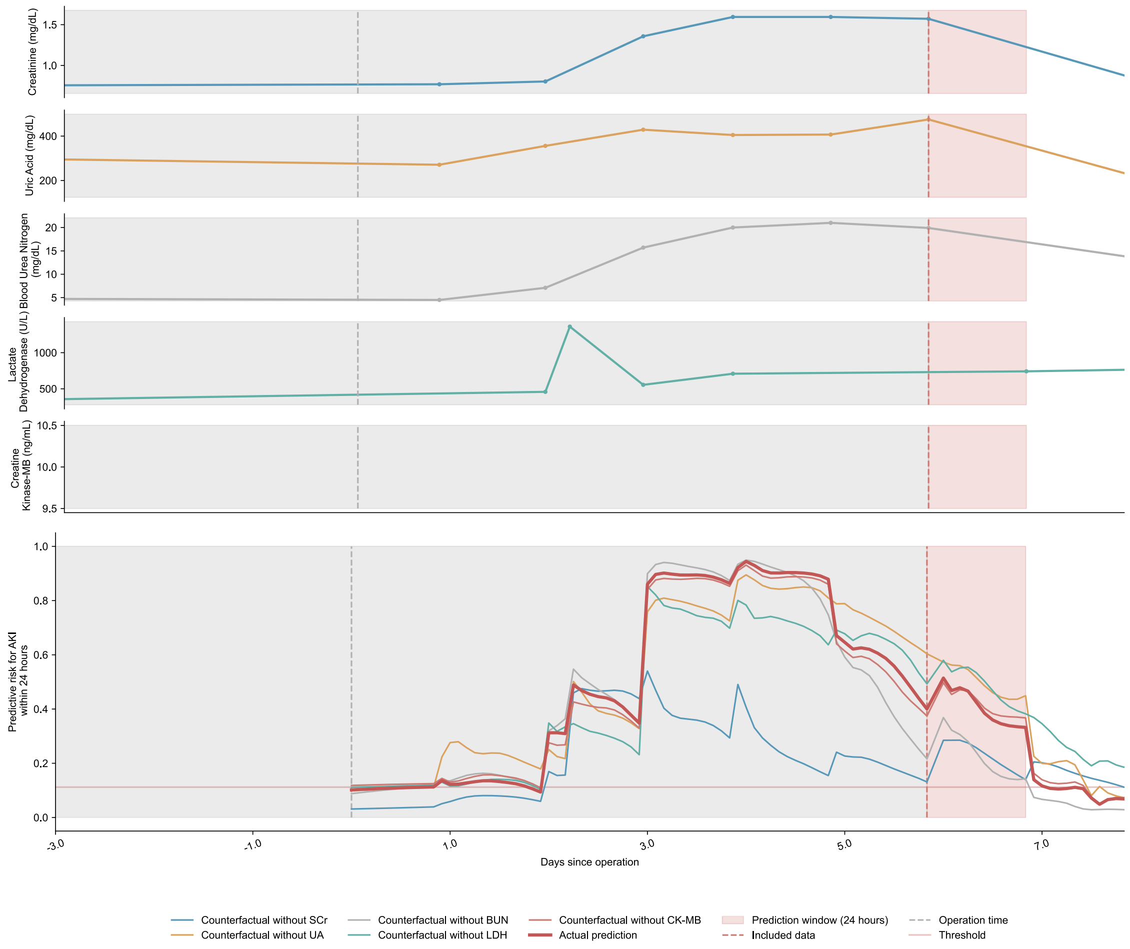


**Supplementary Figure U-3.** A female in their 60s who had undergone coronary-artery bypass grafting alone had respiratory comorbidities. All laboratory examination indicators were stable prior to surgery, and there was no underlying renal disease. On the 6th day after surgery, creatinine, urea nitrogen, uric acid and lactate dehydrogenase began to change unsteadily and eventually CSA-AKI occurred.

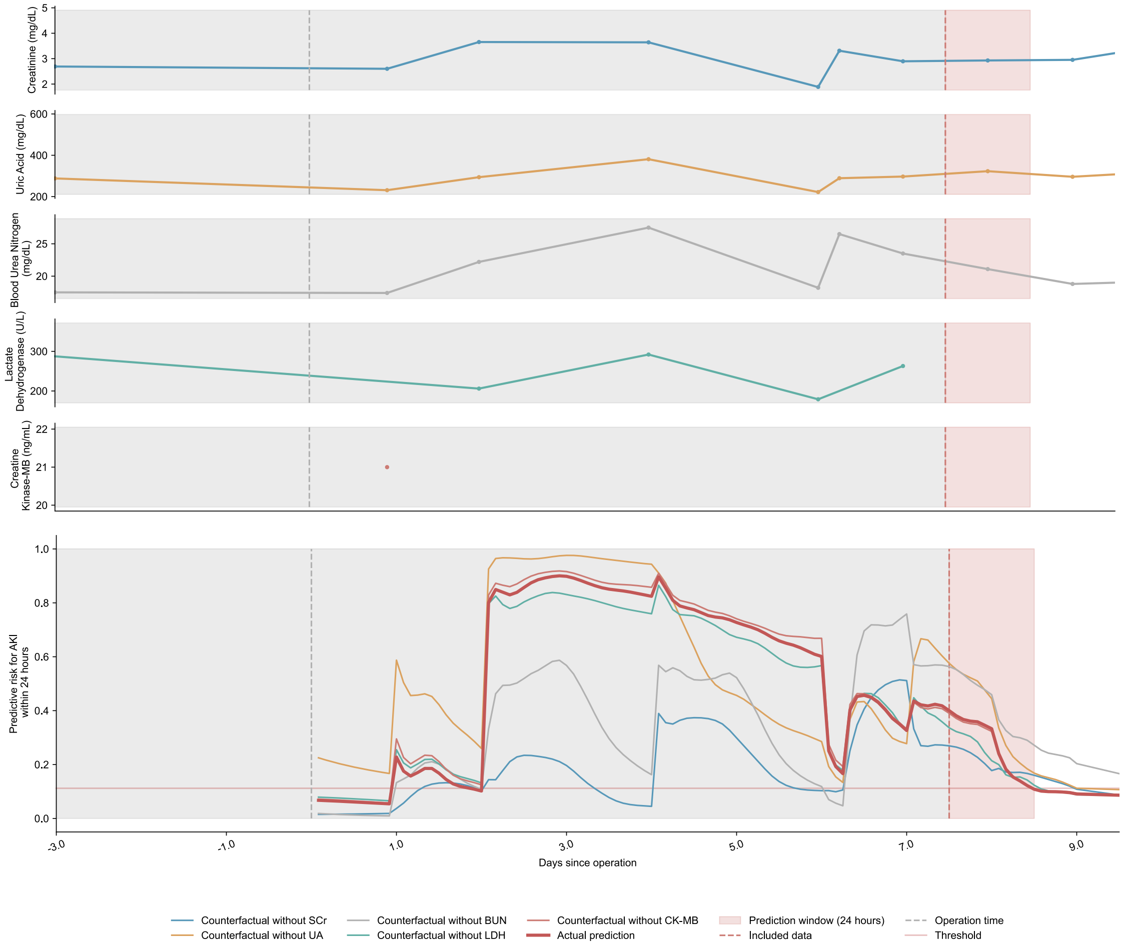


**Supplementary Figure U-4.** A male in their 60s with multiple comorbidities (including chronic heart failure, hypertension, diabetes mellitus, chronic kidney disease, chronic obstructive pulmonary disease) underwent coronary-artery bypass grafting. Creatinine, urea nitrogen, uric acid, and lactate dehydrogenase increased and decreased with time after surgery, but the creatine kinase enzyme was measured only once after surgery. The patient eventually developed CSA-AKI.

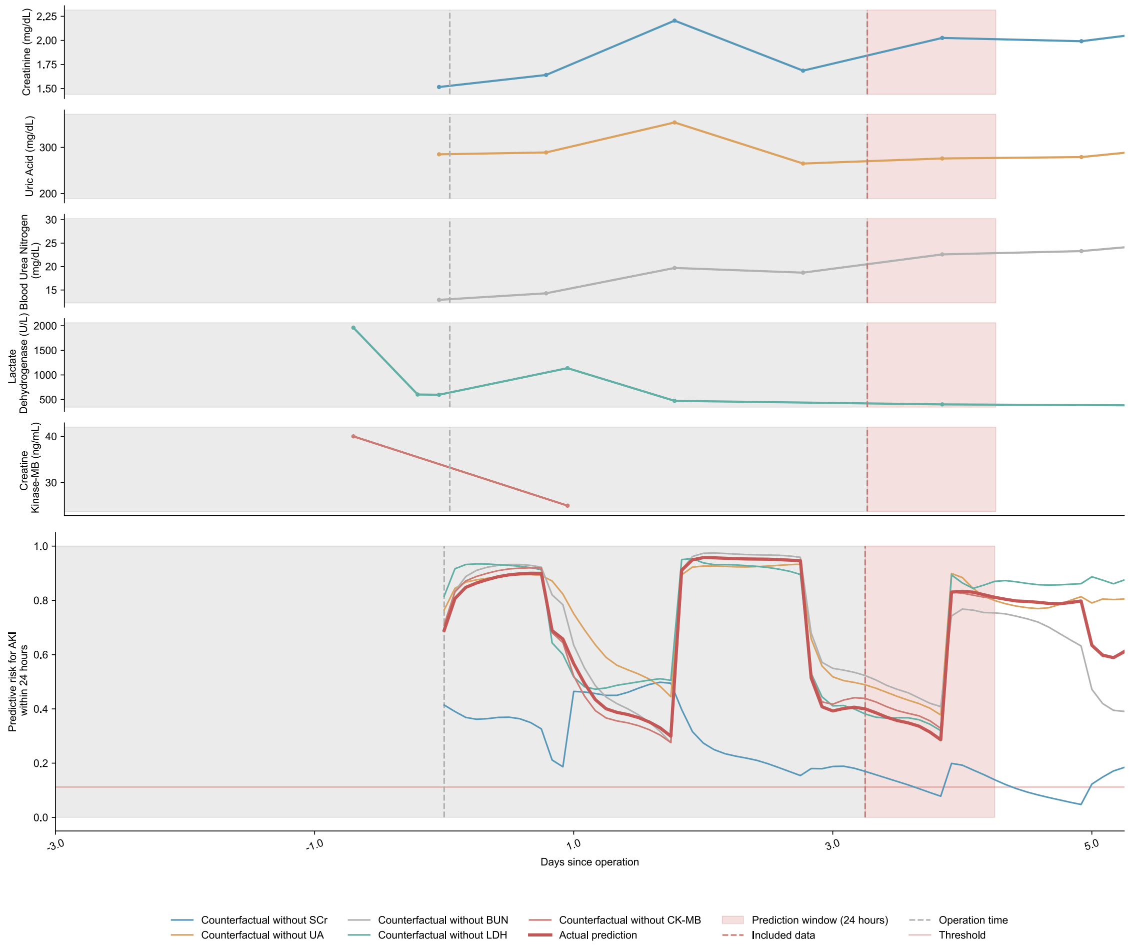


**Supplementary Figure U-5.** A male in their 70s with diabetes mellitus, who was urgently admitted for coronary artery bypass grafting. Due to the emergency surgery, the pre-surgery laboratory examination was measured only once. After the surgery, the creatinine, urea nitrogen, uric acid, lactate dehydrogenase, and creatine kinase enzymes significantly changed over time, and the patient eventually developed CSA-AKI.
